# The etiology of mandibular anterior arch collapse and mesial molar drift: A preliminary study

**DOI:** 10.64898/2026.06.25.26356639

**Authors:** Elaine C. Boosalis Toaddy, Steven D. Marshall, Emma Mueldener, James C. Thomas, Kendra Bogert-Baird, Thomas E. Southard, Kyungsup Shin

## Abstract

Relapse of aligned mandibular anterior teeth and the progressive collapse of the mandibular anterior arch are historically striking problems for orthodontists. The etiology of this collapse, and the cause of mesial molar drift, are unknown. However, light continuous (quasi-continuous) intra-oral pressures and forces applied to the mandibular dentition have been implicated. To explore this further, we use three-dimensional finite element analysis to investigate the influence of these intra-oral loads (tongue pressure, lip-cheek pressure, and interdental force) on mandibular arch collapse and mesial molar drift. Dentitions of three-dimensional finite element mandibular models were subjected to a wide range of simulated tongue pressures, lip-cheek pressures, and transseptal fiber-mediated interdental forces reported in the literature. Resulting crown displacement measurements from these isolated loads were made along with measurements resulting from simultaneous combined application of literature-defined mean tongue pressure, lip-cheek pressure, and interdental force. Our results indicate that tongue pressure alone results in generalized arch expansion and tooth spacing while lip-cheek pressure and interdental force result in generalized arch collapse, anterior crowding, and mesial molar displacement. Simultaneous application of tongue pressure, lip-cheek pressure, and interdental force mean values, as would occur in vivo, results in incisor crowding, intercanine width reduction, and mesial molar displacement. Our results suggest mandibular anterior arch collapse (incisor crowding / intercanine width reduction), and mesial molar displacement result from simultaneous application of tongue pressure, lip-cheek pressure, and interdental force.

## Introduction

Relapse of aligned mandibular anterior teeth and the progressive collapse of mandibular intercanine width remain significant, persistent, and perplexing challenges for orthodontists. Ideally, tooth positions improved by orthodontic treatment would be maintained in a state of equilibrium throughout the remainder of the patient’s life - obviating the need for mechanical retention. However, this is not reality. Instead, to maintain orthodontic tooth alignment post-treatment, life-long retention is generally required.

Historical and longitudinal research, beginning with case reports by Milo Hellman who lamented, “we are in almost complete ignorance of the specific factors causing relapse and failure.” [1], and followed by seminal works of Little [2], Sinclair [3], and Shapiro [4], has consistently demonstrated that mandibular anterior crowding tends to worsen over time and that intercanine width decreases over time, regardless of whether patients undergo extraction treatment, non-extraction treatment, or even remain untreated. The underlying causes of these phenomena are considered complex and multifactorial, but tooth positions are suggested to be the product of an equilibrium between mechanical (occlusal-related), periodontal, and soft-tissue forces. Unfortunately, our understanding of the biologic factors leading to position equilibrium, or position imbalance and relapse, is incomplete.

The equilibrium theory of tooth position has been defined as teeth occupying a stable position resulting from the pressures of opposing forces.[5] Although other factors may play a role (increasing overbite, continued mandibular growth, or the anterior component of occlusal force), Proffit suggested that the major *primary* factors affecting dental equilibrium (or position imbalance and relapse) appear to be resting pressures of tongue and lips plus forces created within the periodontal tissues.[6]

Resting tongue pressure, resting lip pressure, and resting cheek pressure have been measured in numerous populations with various technologies.[7–19] Influenced by methodology, anatomical site, population, and sample size, values reported for these forces vary widely. Previously reported resting tongue pressure values range from 0-22 kg/m^2^ to 135-150 kg/m^2^ (Fig 1).[7,10,11,12,15,19] Previously reported resting lip-cheek pressure values range from 0-30 kg/m^2^ to 270-450 kg/m^2^ (Fig 2).[8,9,11,13,14,16,17,18]

**Fig 1.**
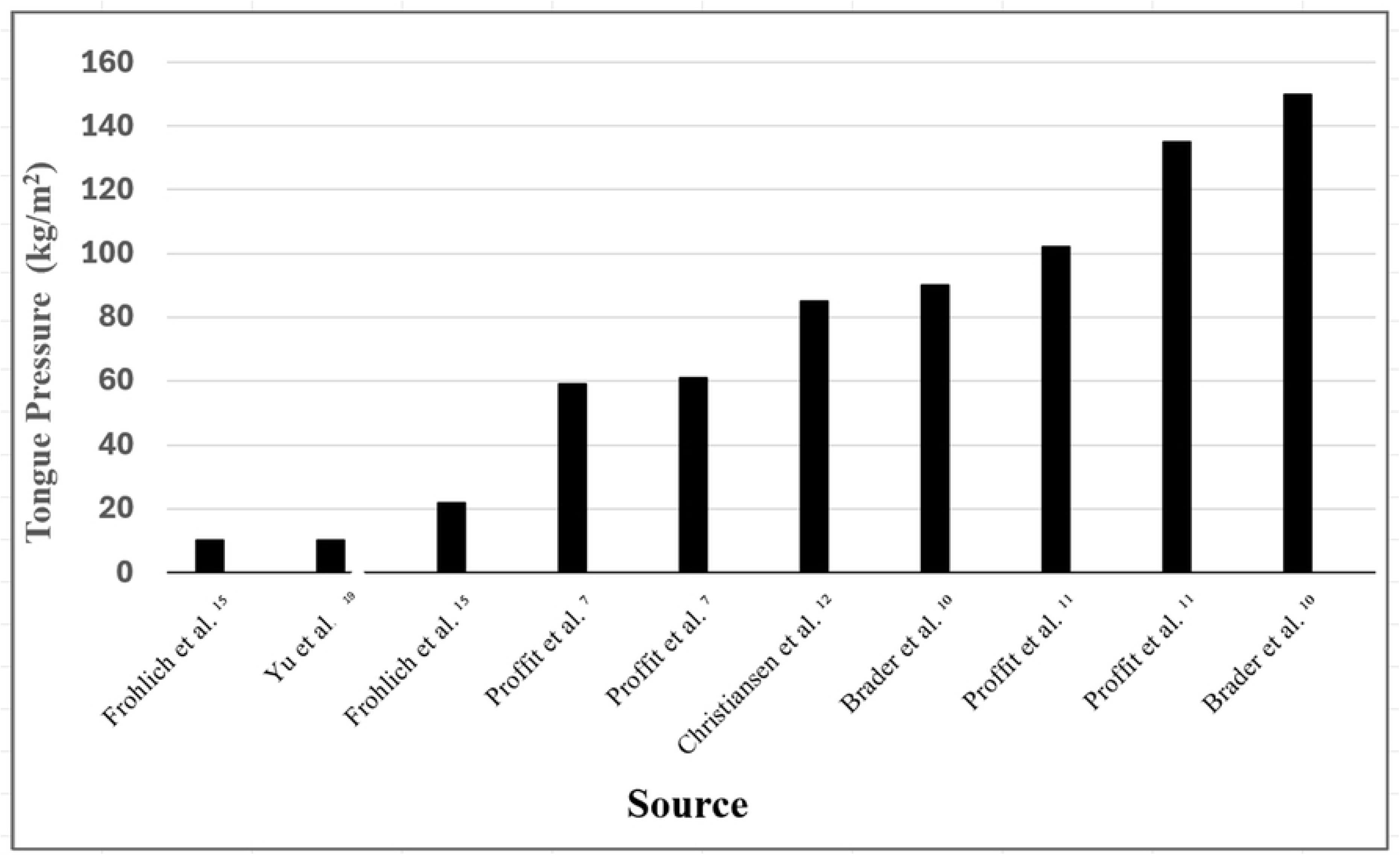
Resting tongue pressure from various studies. Tongue pressures (kg/m^2^) reported from various studies.

**Fig 2.**
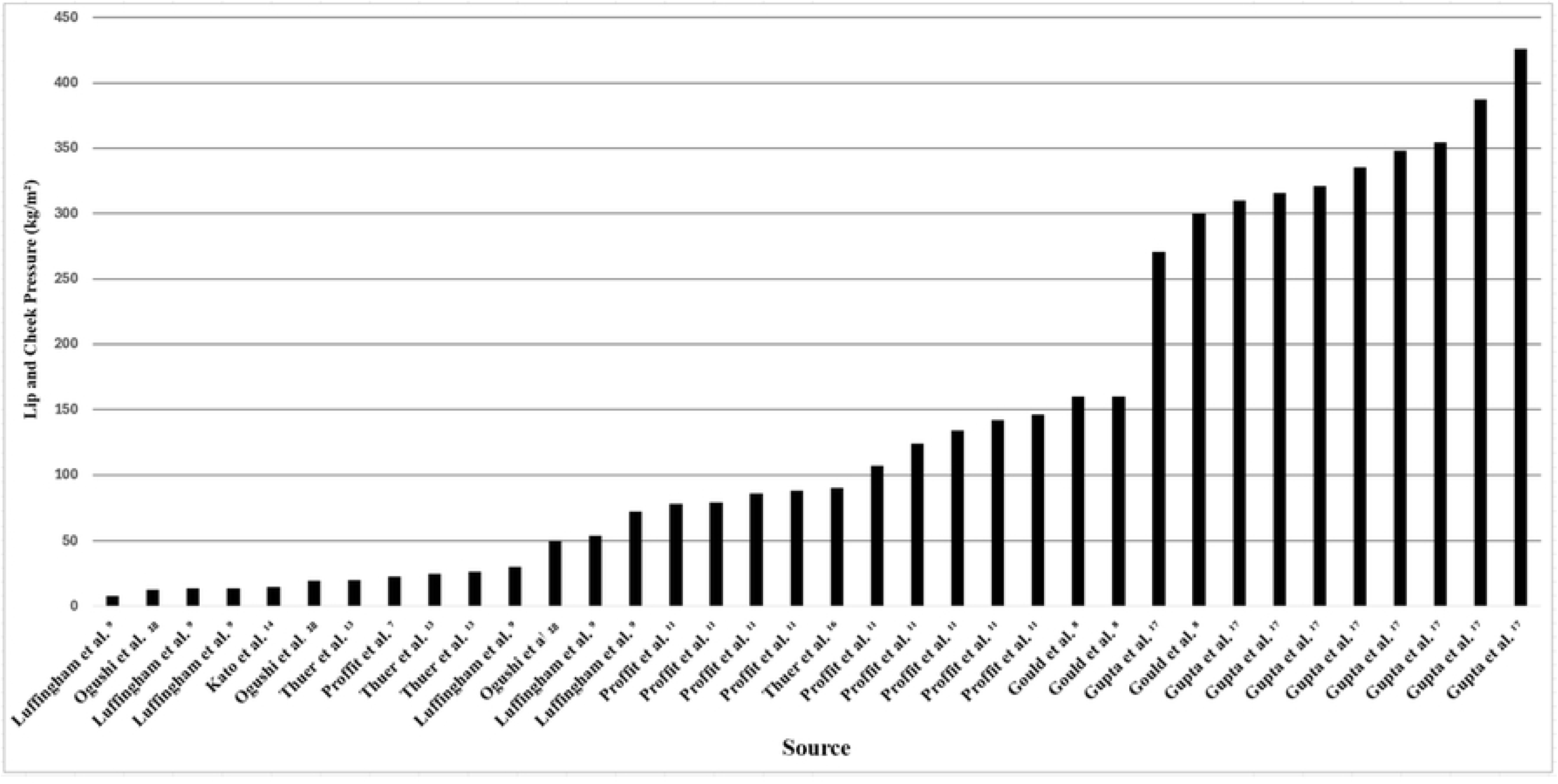
Resting lip and cheek pressure from various studies. Resting lip and cheek pressures (kg/m^2^) reported from various studies.

In addition to tongue and lip-cheek pressure, others have speculated that a third continuous, or quasi-continuous, intra-oral force may affect tooth position equilibrium / imbalance - the interdental force mediated by gingival transseptal fibers.[20–24] Transseptal fibers originate and insert from tooth to bone and from tooth to tooth.[20] Fibers attaching to bone may remodel, but those inserting into neighboring teeth may lack remodeling capacity, and Aisenberg suggested that this inability to remodel could subject the mandibular anterior segment to collapse and relapse.[20] By repeatedly removing posterior contacts as they re-established in *Macaca irus* monkeys, Moss and Picton [21] found that teeth continued to migrate, leading the authors to conclude that interdental (transseptal fiber) forces likely contribute to these movements. Edwards highlighted the retention challenge posed by transseptal and supracrestal fibers noting their impact on rotational movements and post-treatment relapse.[22] Following bilateral premolar extractions in *Macaca rhesus* monkeys, and surgically severing transseptal fibers unilaterally after space closure, Parker demonstrated that intact (non-surgical) sides experienced migration-relapse while surgical sides maintained stability, and concluded that interdental (transseptal fiber) forces contribute, in part, to mandibular incisor relapse.[23] In the only quantitative measurement of interdental force to date, Southard [24] studied interdental force values prior to and after chewing. They reported that the periodontium exerts a continuous compressive force on the dentition and hypothesized that this force may accelerate mandibular anterior arch collapse.[24] Overall, pre-chewing (resting) interdental force values range from a low of 0-25 g to a high of 60-86.9 g, with a mean of approximately 30-55 g.

Mesial molar drift is another well-recognized phenomenon whose etiology is not fully understood. Some have suggested that it is dependent on growth changes in the lower face and possibly the interdental (transseptal) force system.[21,25,26] Moss and Picton found that mesial drift occurred in *Macaca Irus* monkeys following removal of cheek and tongue pressures and concluded that interdental fibers played a key role in mesial drift.[27] In a follow-up study, interdental sites where fibers were surgically removed in monkeys closed minimally compared to intact fiber sites which closed predictably, leading the authors to conclude that transseptal fibers may be a principal driver in mesial drift.[28]

In summary, progressive mandibular anterior crowding and intercanine width reduction are extensively documented, persistent, and unpredictable phenomena occurring in untreated and orthodontically treated populations regardless of initial occlusion type, extraction regimen, or treatment biomechanics. Numerous factors have been implicated as possible etiologies of mandibular anterior arch collapse with continuous, or quasi-continuous, tongue pressure, lip-cheek pressure, and inter-dental force the primary suspected causes. However, despite decades of speculation, the literature remains fragmented and lacking in terms of studies demonstrating a causal relationship between these loads and mandibular anterior arch collapse probably because these loads are very light and applied over decades – making experimental design exceptionally challenging if not impossible. Clinically, this uncertainty hampers our ability to predict stability, individualize retention protocols, and prevent relapse from occurring in the absence of long-term mechanical retention.

The purpose of this preliminary investigation is to explore the influence of tongue pressure, lip-cheek pressure, and interdental force, individually and in combination, on mandibular anterior arch collapse, incisor crowding, and mesial molar drift using finite element analysis (FEA) modeling. It seeks to address a critical, unexplored knowledge gap in the areas of orthodontic relapse and retention. By scoping this area and identifying teeth displacement trends under pressures and forces implicated in orthodontic relapse, we hope to establish the necessary foundation for future studies and encourage researchers to pursue this line of inquiry.

## Materials and Methods

A virtual mandible and mandibular teeth were imported from the University of Dundee, School of Dentistry.[29] The model was modified to exclude the interior mesh volume of the mental foramina and converted to STL file format. Mandibular resolution was reduced by 60% which lowered the polygon count to 367,174 faces. All parts were imported in STEP (.stp) format into Abaqus/CAE (Dassault Systèmes, Providence, RI) to generate a 3D finite element model using the AP203 Configuration Controlled Design export schema.

According to the theory of orthodontic tooth movement proposed by Schwarz, pressure side tooth loading compresses the periodontal ligament (PDL) and disturbs blood flow.[30] Cell death (hyalinization) results, forming a temporary barrier to root movement until the necrotic tissue is removed by macrophages and until osteoclasts recruited from adjacent bone marrow spaces have resorbed the adjacent bone.[30–35] Studies have shown that this process occurs with forces as low as 0.05N.[36] To simplify FEA study results, bone and other tissues are most often modeled as responding linearly despite evidence suggesting a non-linear, time dependent tissue response during tooth movement (e.g. initial phase, lag phase, post-lag phase), and the periodontal ligament can be included as part of the model or teeth may be connected directly to bone.[37–41] In selecting material properties for orthodontic FEA studies, Romanyk emphasized the need to select values that align with study goals.[41] For example, Cattaneo considered three different Young’s moduli for bone (ranging from full cortical bone to bone marrow) but chose to use the mean of the three for their model.[42] Bone has even been modeled as a rigid body.[43] If bone is assumed to be linearly elastic, homogeneous, and isotropic, the value of Young’s modulus chosen will influence FEA model part strain (proportional to stress), and part displacement, under load. In the present study, mandibular bone was assumed to be isotropic, homogeneous, and linearly elastic.

Teeth were converted into discrete rigid shells to reduce computation time and stay within our Academic Licensing limit restriction (250,000 nodes). The dentition was aligned in the mandible with incisal edges and cusp tips lying along the X-Z plane. Dental sockets were cut into the mandible precisely matching teeth roots which were tie-constrained to trabecular (cancellous) bone with a Young’s modulus of 6 kPa and a Poisson’s ratio of 0.3.[44, 45] The mandible was meshed with tetrahedral elements (193,000 total elements) suitable for irregular geometries (Fig 3). Shell teeth were meshed with triangular elements (81,000 total elements). Fixed boundary conditions were established for elements at the base of the mandible, distal to the second molars, and at the condyles to prevent model body motion when loads were applied. This arrangement established the pre-load, or undeformed, baseline assembly used in all models.

**Fig 3.**
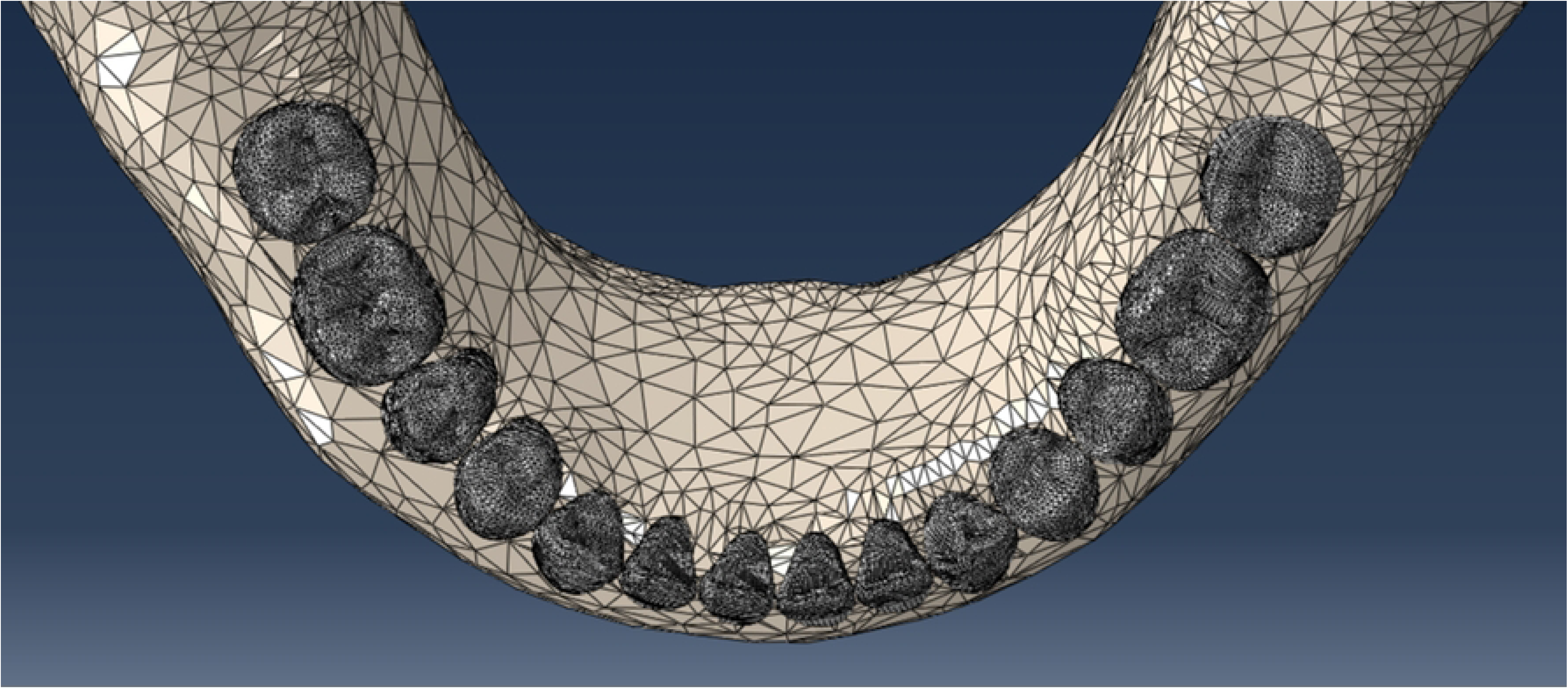
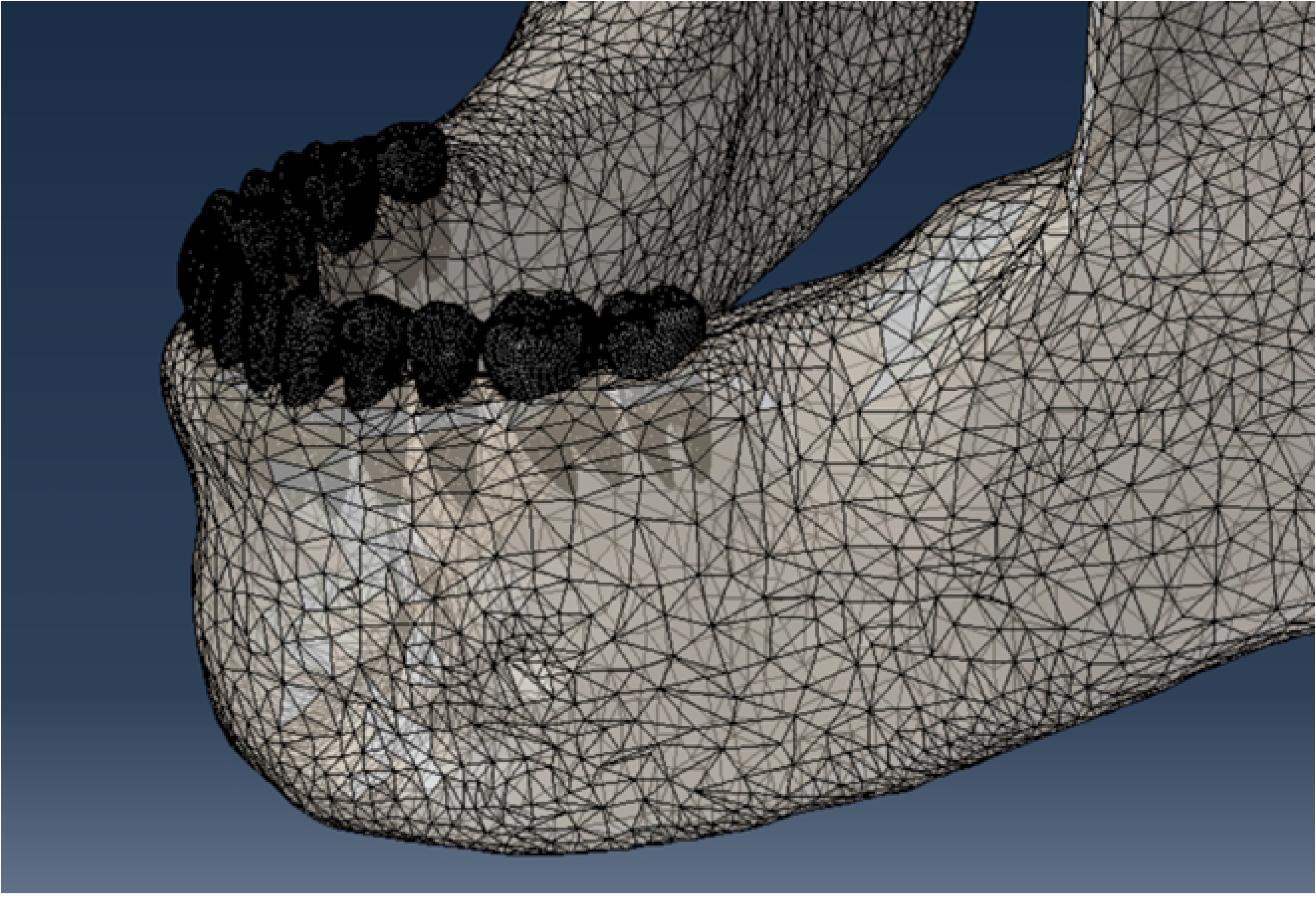
Mandibular meshed model (A, occlusal view; B perspective view).

### Model Loading

Directional vectors were calculated in the X-Z plane interdentally between centroids of adjacent teeth, toward the lingual perpendicular to the arch, and toward the labial perpendicular to the arch (Fig 4).

**Fig 4.**
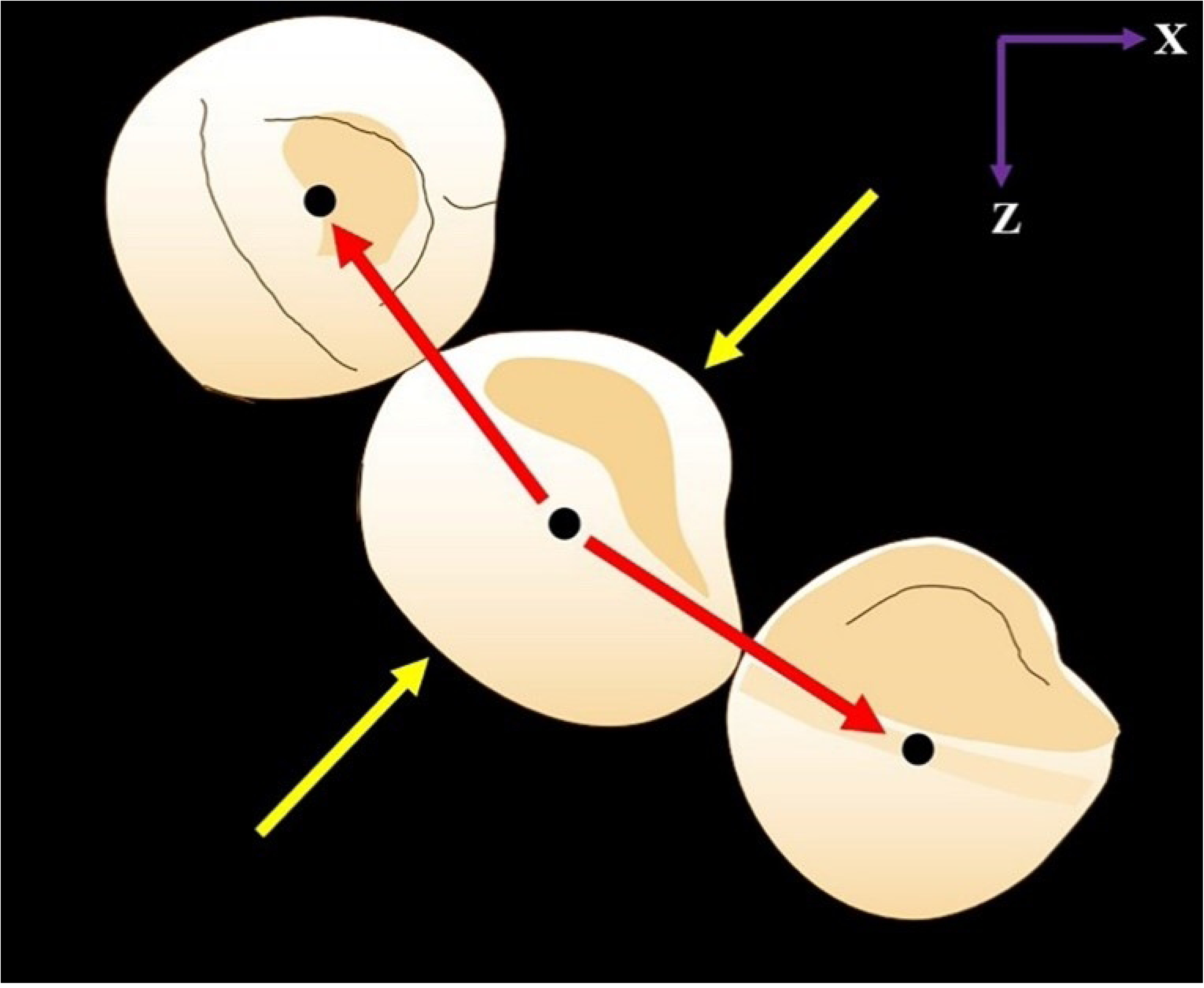
Sample (mandibular right canine) load directional vectors. Interdental vectors (red arrows) between canine-lateral incisor centroids and between canine-first premolar centroids. Labial and lingual directional vectors perpendicular to the arch (yellow arrows).

Dentition positional data, tongue pressure data (low range value, mean value, high range value), lip-cheek pressure data (low range value, mean value, high range value), and interdental force data (low range value, mean value, high range value) were entered in a spreadsheet (Microsoft Excel) and used to calculate directional vectors and F_x_ and F_z_ force components applied to teeth crowns for each of nine models (three tongue pressure models, three lip-cheek pressure models, three interdental force models).

Labial / lingual crown surface areas [46], lip-cheek pressures, and labial / lingual directional vectors were used to calculate lip-cheek F_x_ and F_z_ force components applied to each tooth crown centroid. Macroscopic crown displacements resulting from three lip-cheek pressure magnitudes were evaluated: 10.4 kilogram per meter squared (kg/m^2^, low range literature value), 136.5 kg/m^2^ (mean literature value), and 328.3 kg/m^2^ (high range literature value). Similarly, macroscopic crown displacements resulting from three tongue pressure magnitudes were evaluated: 10 kg/m^2^ (low range literature value), 100 kg/m^2^ (mean literature value), and 150 kg/m^2^ (high range literature value). Geometric nonlinearity and frictionless interaction between contacting teeth crowns was incorporated into all loading analyses.

The interdental (transeptal fiber) force magnitude, and interdental directional vectors, were used to calculate the interdental force F_x_ and F_z_ force components applied to each tooth crown centroid. Macroscopic crown displacements resulting from three interdental force magnitudes were evaluated: 0.0060 kg (low range literature value), 0.0366 kg (mean literature value), and .0869 kg (high range literature value).

A final simultaneous loading of the mandibular dentition with all three mean intraoral loads was performed. Macroscopic crown displacements resulting from dentition loading of 100 kg/m^2^ (mean value) tongue pressure, 136.5 kg/m^2^ (mean value) lip-cheek pressure, and 0.0366 kg (mean value) interdental force were evaluated.

### Model Measurements

Teeth positions were analyzed before and after tongue pressure, lip-cheek pressure, or interdental force were applied individually for all nine simulations to determine teeth displacements (arch changes) due to pressure/force loading. Consistent reference nodes were selected as anatomic dental landmarks between models to provide precise displacement measurements which included intermolar width, intercanine width, arch perimeter, anterior arch perimeter, and arch length (Figs 5 and 6).[47]

**Fig 5.**
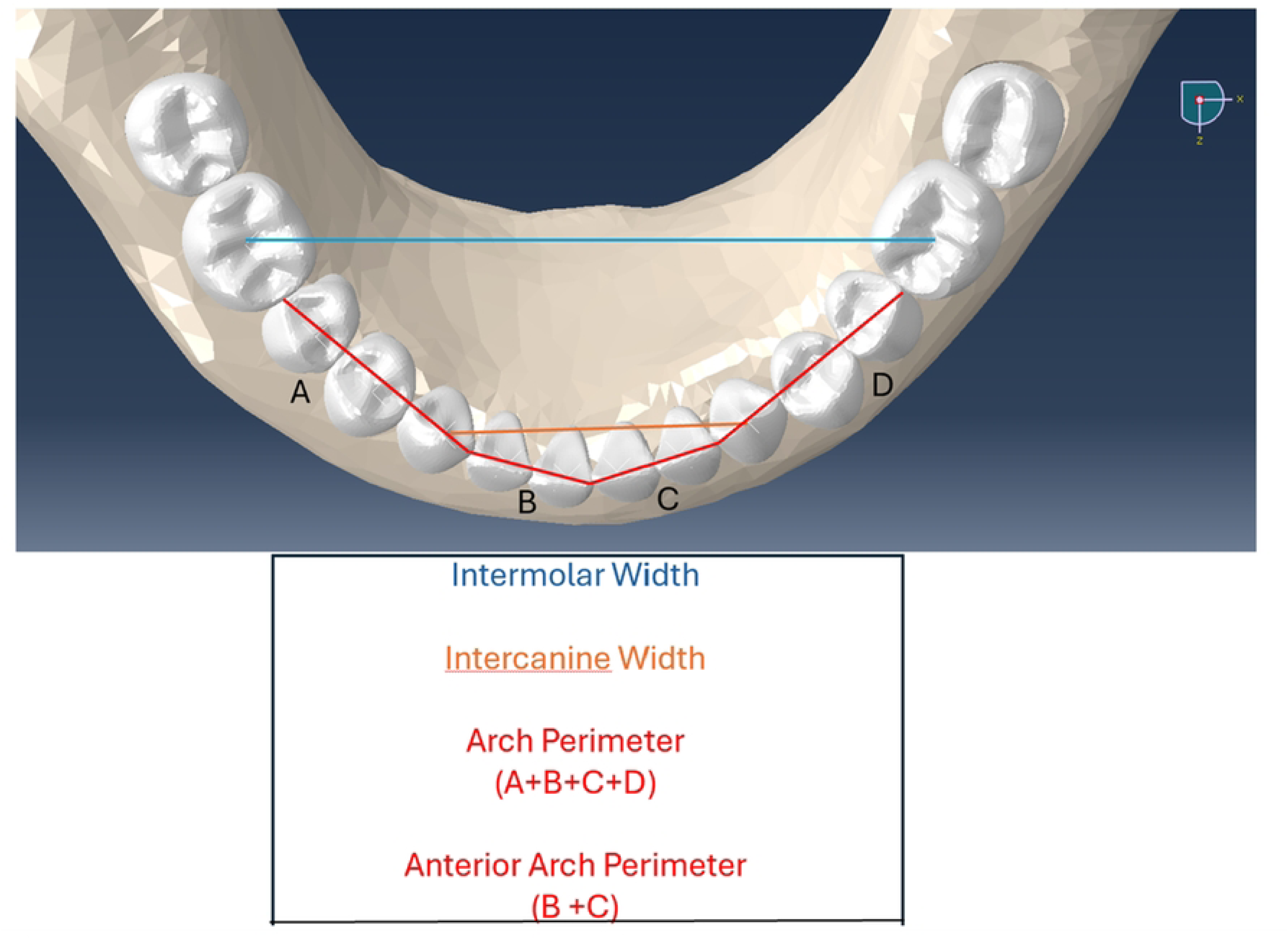
Measurement of intermolar width, intercanine width, arch perimeter, and anterior arch perimeter. Intermolar width (blue). Intercanine width (red). Molar-to-molar arch perimeter = A+B+C+D. Canine to canine arch perimeter = A+B.

**Fig 6.**
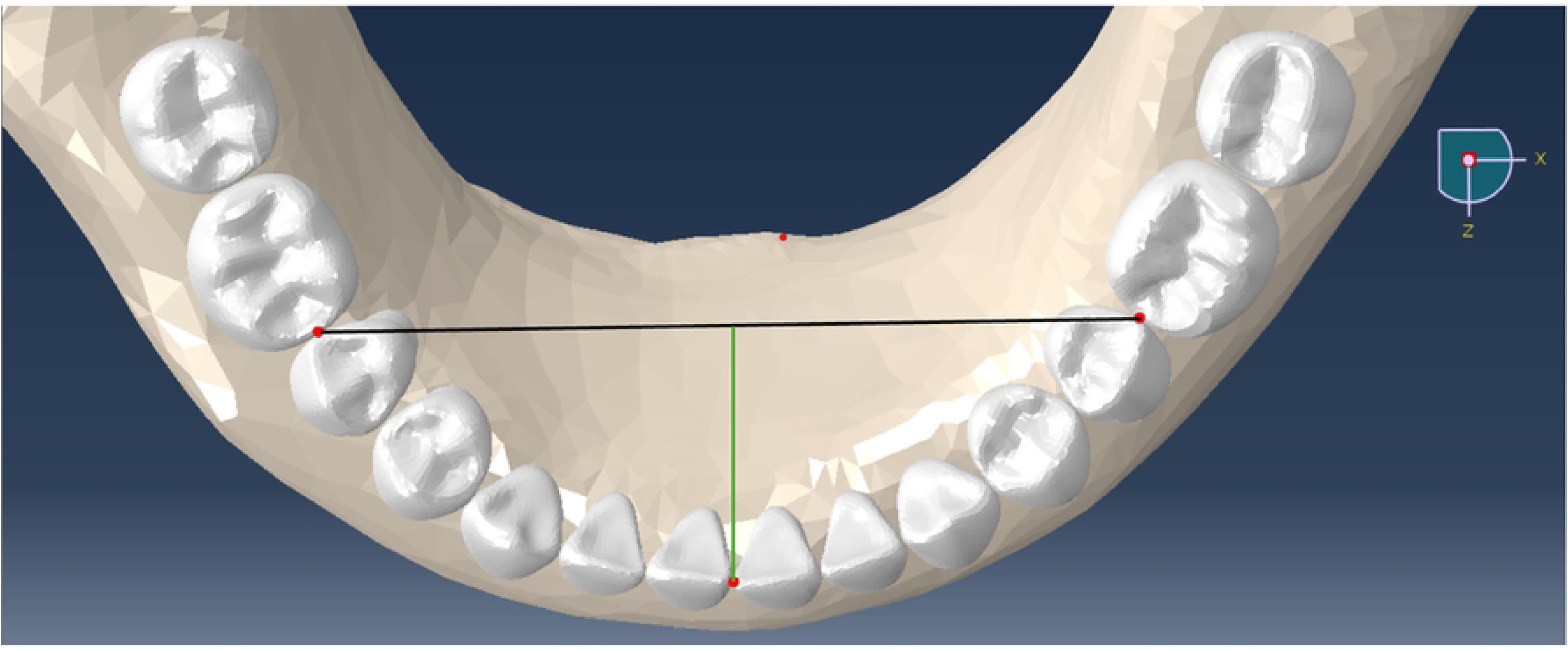
Measurement of mandibular arch length. Mandibular arch length (green line) measured as shortest (perpendicular) distance between consistent reference node on the left central incisor mesial (red dot) and line between first molar mesial reference nodes at contact points.

Changes in mandibular incisor crowding (or spacing) were quantified by subtracting the sum of the four incisor mesiodistal widths from the anterior arch perimeter before, and after, loading. Positive differences indicated increased spacing with loading. Negative differences indicated increased crowding with loading. Note: a slight amount of incisor spacing (1.06 mm) was initially present in all identical, unloaded models.

Scaled crown displacement vector plots (Fig 7) were created for each model to provide a qualitative interpretation and comparison of relative crown movements when subjected to tongue, lip-cheek, or interdental force loads. The model exhibiting the lowest maximum displacement was accepted as baseline using the Abaqus default scale setting. For all other models, displacement vector lengths were scaled proportionally based on their respective maximum displacement values thus allowing for visually meaningful comparisons between pressure, or force, effects. Vector color coding indicated directional displacement with red vectors representing mandibular arch collapse tendency and yellow vectors indicating mandibular arch expansion tendency. Partial translucency of model assembly aided vector visualization.

**Fig 7.**
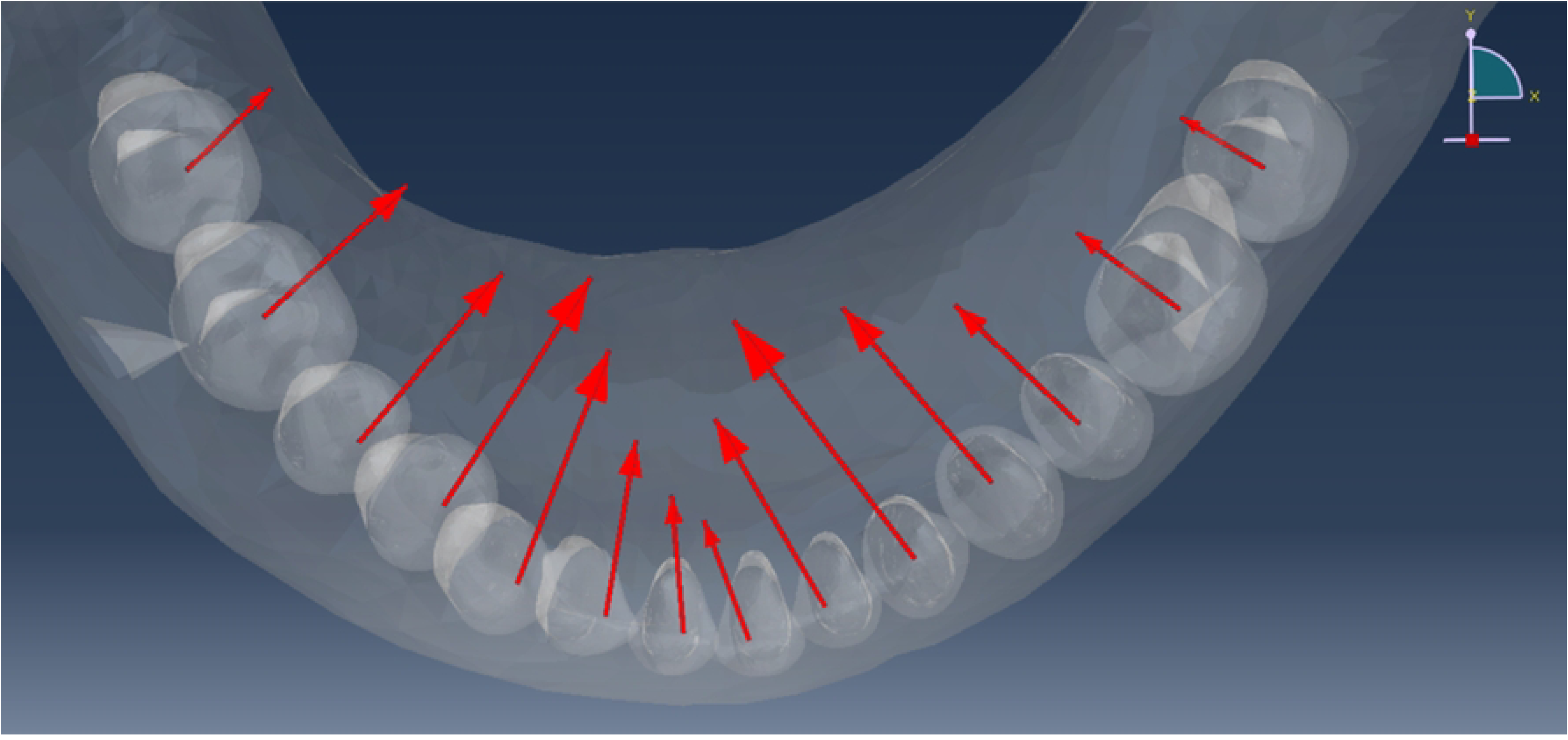
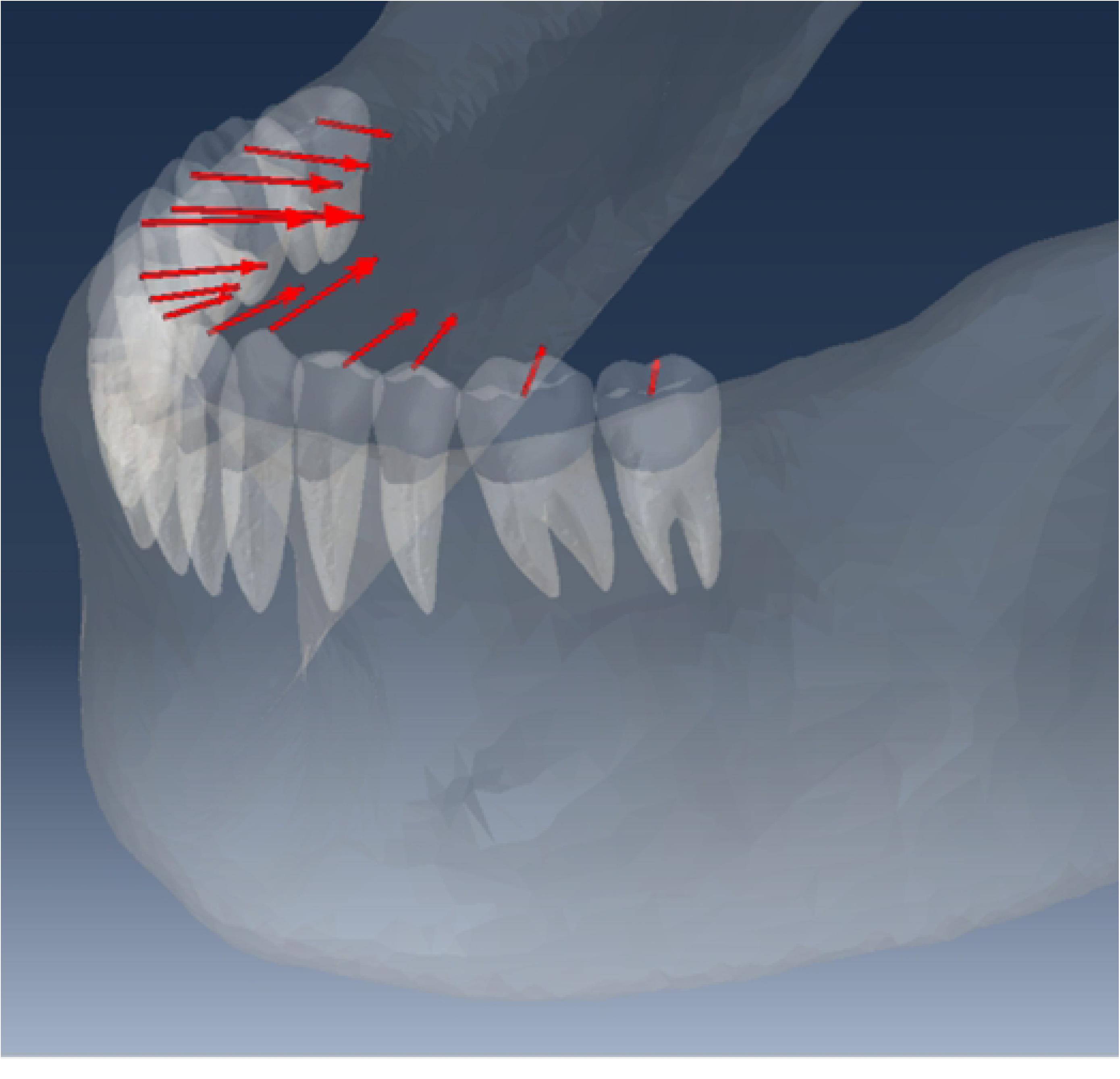
Example of scaled displacement vector plots. (7**A**, occlusal view; 7**B** perspective view). Vector lengths indicate relative crown displacement magnitudes in directions shown. Red vector color indicates crown displacements tending to collapse mandibular dental arch.

Additionally, to reflect conditions found intra-orally, a mandibular arch was subjected to *simultaneous* combined loading by tongue, lip-cheek, and interdental force using the mean values described in the literature: 100 kg/m^2^ (mean value) for tongue pressure, 136.5 kg/m^2^ (mean value) for lip-cheek pressure, and 0.0366 kg (mean value) for interdental force. For this simultaneous loading, the measurements previously described were made along with a superimposition of unloaded and loaded models to provide a further qualitative interpretation of crown displacements.

### Model Verification and Statistical Analysis

Verification included re-running models at greater, and lesser, mesh densities to determine whether this would affect measurement results. We re-meshed and ran our mean tongue pressure model using a mesh ten percent denser than our original (baseline) mesh. We calculated the resulting differences in measurement values between the denser mesh and baseline mesh models. This comparison was repeated using a mesh ten percent less dense than our original.

The strength of linear association between each tongue pressure, lip-cheek pressure, and interdental force loading magnitude and the resulting mandibular arch measurements were calculated with Pearson’s correlation coefficient. To quantify the uncertainty of these measurements, confidence intervals were also determined.[47] Due to the small sample size of each independent and dependent variable pairing, t intervals were used in place of the typical Z approximation.[48]

## Results and Discussion

### Tongue Pressure Effects on Dental Displacement Patterns

A representative displacement vector plot of mandibular teeth resulting from a tongue pressure of 100 kg/m^2^ (mean literature value) is shown in Fig 8. As illustrated, tongue pressure creates generalized outward, expansive, crown movement. Interestingly, the greatest displacements from tongue pressure are found at the canines and premolars, and a *mesial* displacement component is found for all crowns as the tongue applies pressure perpendicular to the arch. Qualitative examination of teeth displacements revealed that tongue pressure resulted in minimal root movement compared to crown movement.

**Fig 8.**
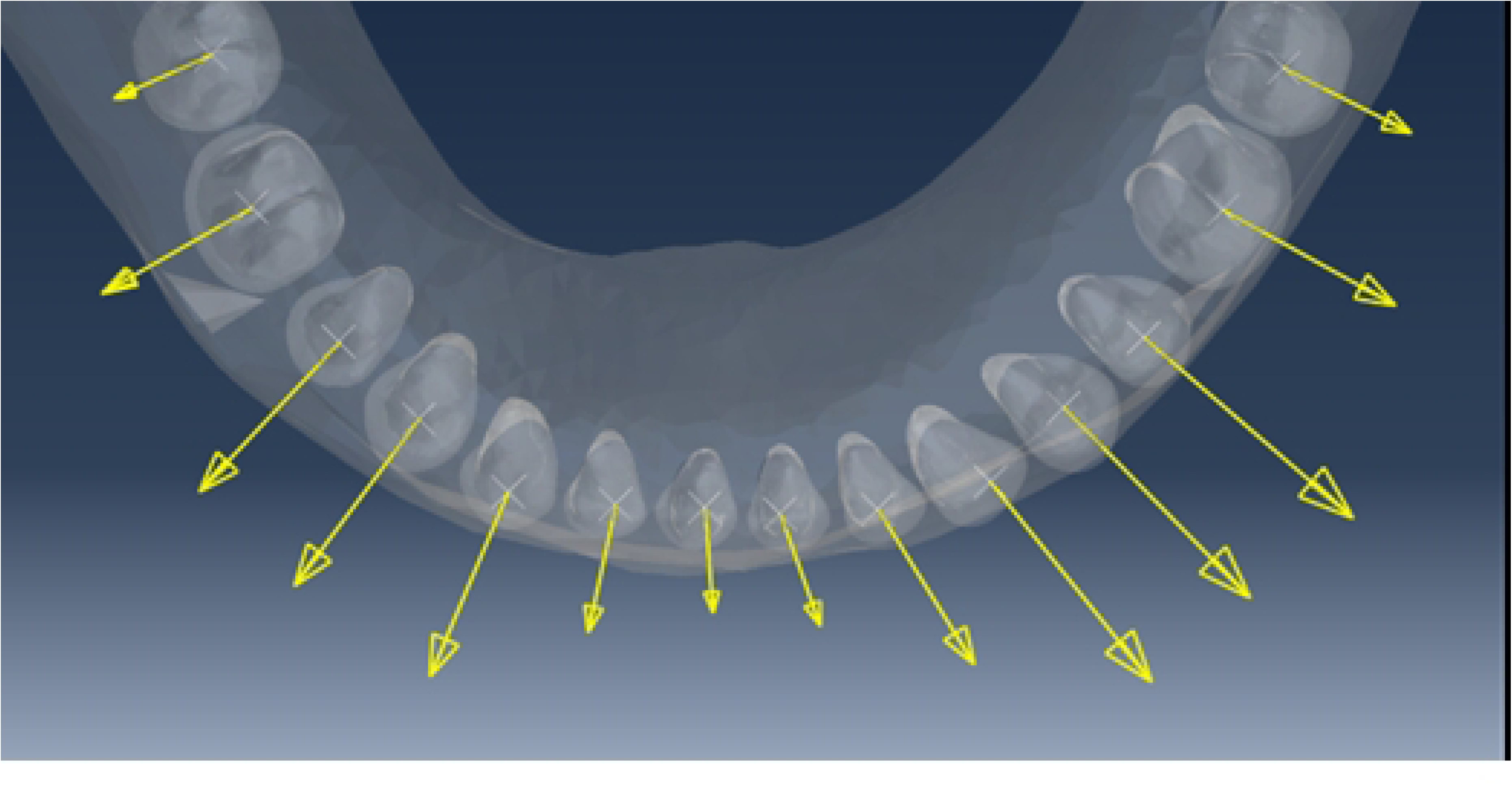
Scaled displacement vector plot of mandibular teeth crowns resulting from tongue pressure of 100 kg/m2 (mean literature value). Yellow vector color indicates crown displacements tending to expand the mandibular dental arch.

Occlusal (X-Z plane) images of the unloaded mandibular arch (Fig 9) and a loaded mandibular arch (Fig 10, 150 kg/m^2^, high range value) are shown. The high range value loaded arch was selected to illustrate the generalized expansive, outward dental displacements caused by tongue pressure. Plots of corresponding arch measurements of unloaded and three loaded mandibular arches are provided in Fig 11. These results quantitatively corroborate the qualitative findings of tongue pressure suggested in the vector plot (Fig 11). That is, tongue pressure results in significant anteroposterior and transverse expansion.

**Fig 9.**
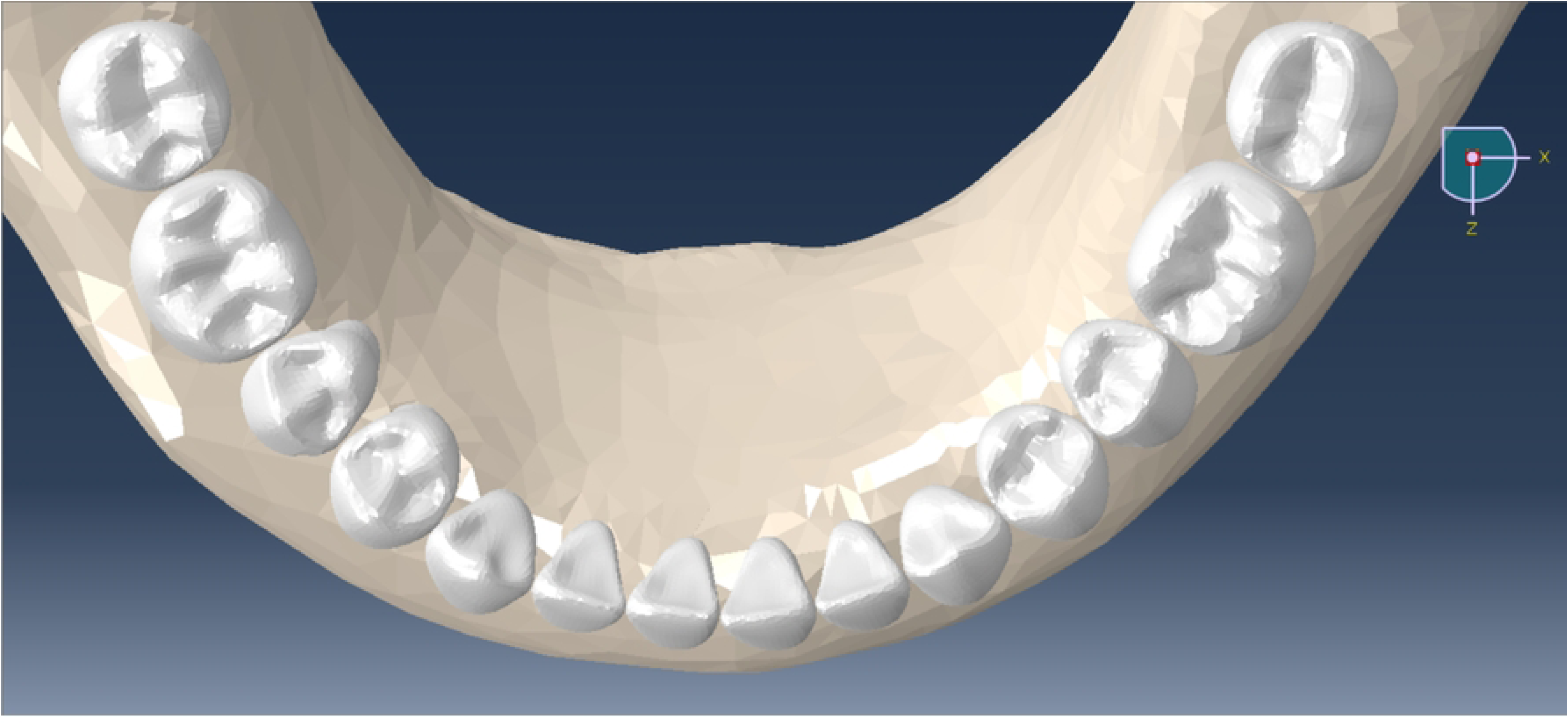
Mandibular arch prior to application of pressures.

**Fig 10.**
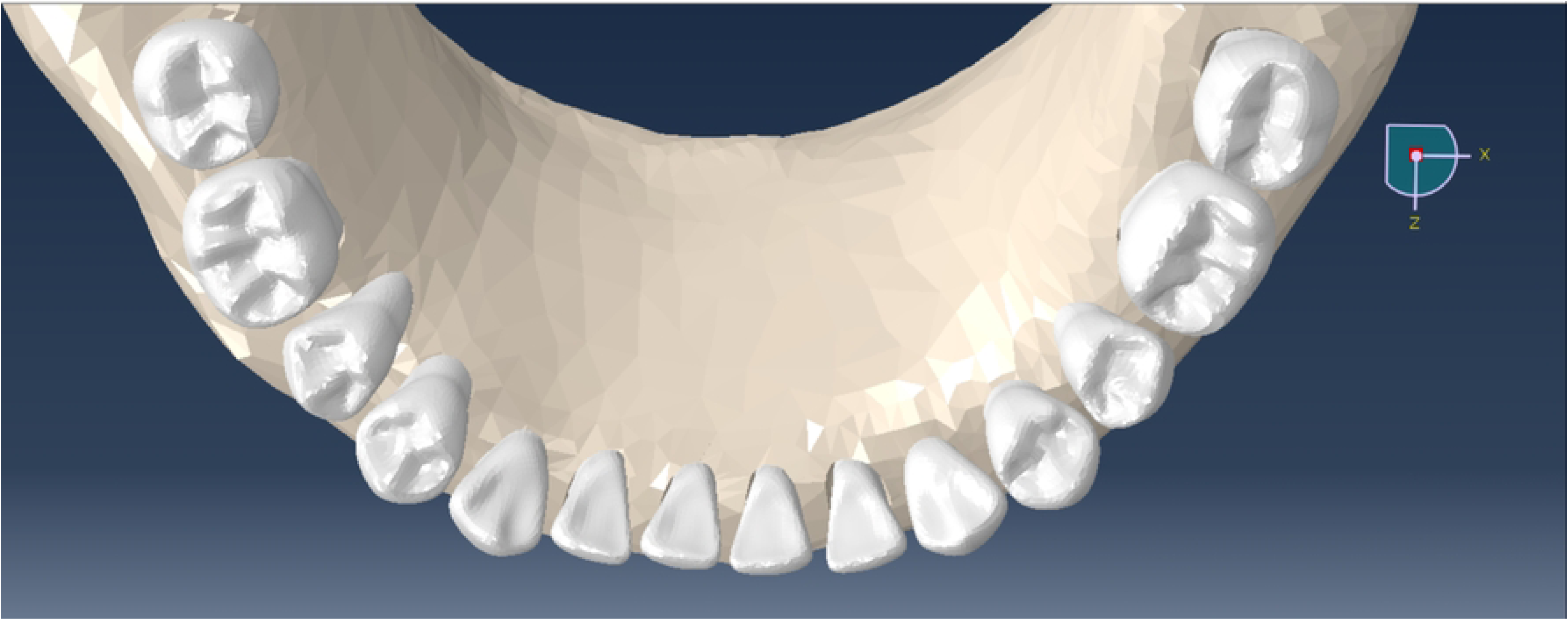
Mandibular arch loaded using tongue pressure model (150 kg/m^2^, high range literature value).

**Fig 11.**
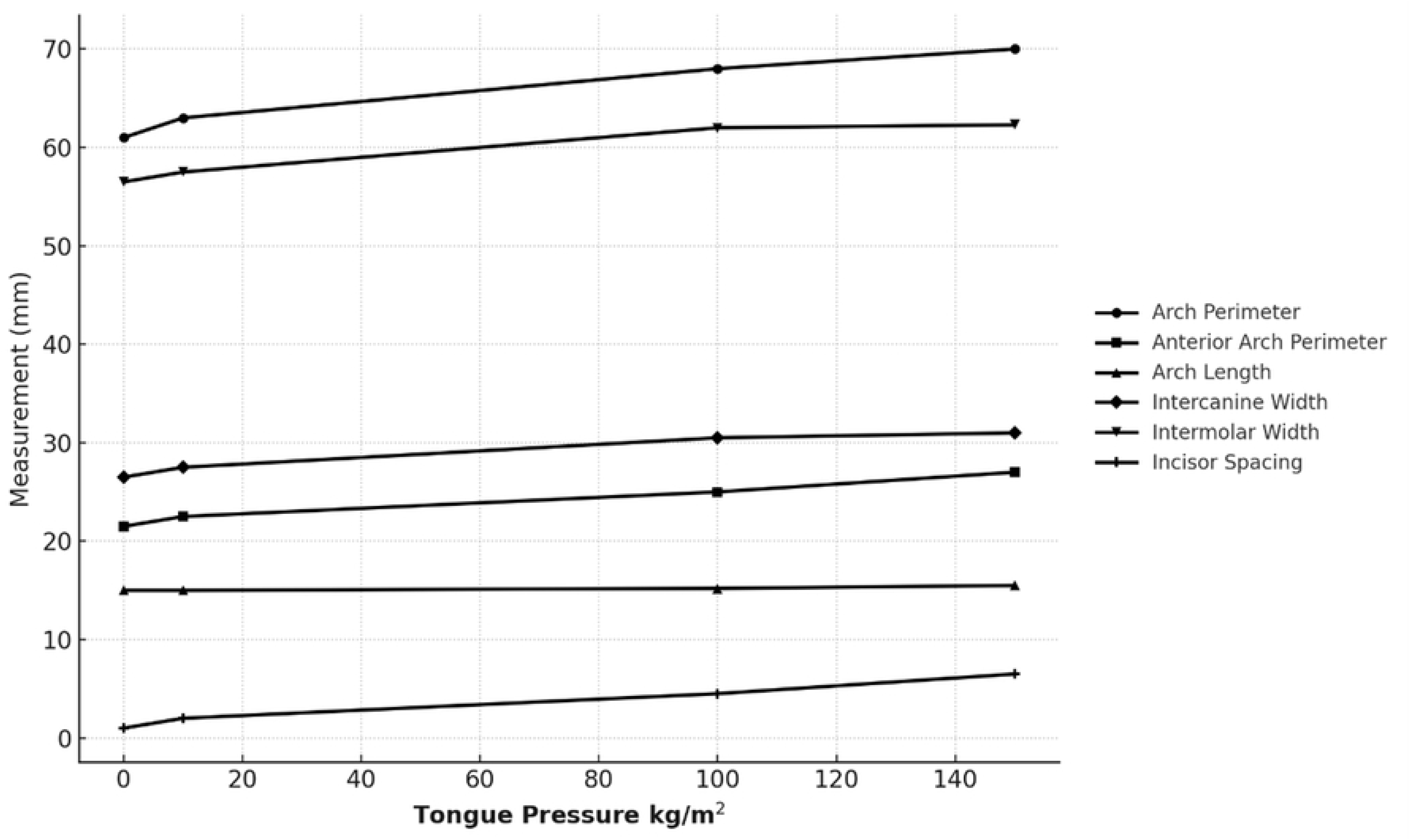
Effect of tongue pressure (0-150 kg/m^2^) on dental arch parameters.

Under the influence of a mean (literature defined) tongue pressure, mandibular arch perimeter increased by 6.7mm, or 11%, and the anterior arch perimeter increased by 3.4mm, or 16%. These changes are visualized as incisor *and* posterior teeth spacing with loading. Surprisingly, even at the high range value of tongue pressure, the arch length increased by only 0.42mm, or 3%. With the amount of incisor labial tipping movement (compare incisor positions in Figs 9 and 10), one would expect a greater corresponding increase in arch length. However, this result is explained by the fact that tongue pressure created slight mesial molar movement which reduced arch length increases - movement confirmed by determining molar reference node positions in unloaded and loaded models relative to mandibular encastre elements.

Intercanine and intermolar widths expanded under the influence of tongue pressure. Under the influence of a mean tongue pressure, the mandibular intercanine width increased by 4.2mm, or 16%, and the intermolar width increased by 5.8mm, or 10%. Minimal additional increase in either intercanine or intermolar widths occurred when the tongue pressure increased from 100 kg/m^2^ to 150 kg/m^2^. A marked increase in incisor spacing (3.4mm) occurred under the influence of mean tongue pressure and continued to increase at the high range tongue pressure value.

Verification results proved excellent. Of 18 loaded model measurements made, 16 were perfect with no difference between baseline mesh and denser mesh, or less dense, models. Only loaded intermolar widths revealed a negligible displacement difference (0.02mm, 0.03%) between baseline mesh and denser, or less dense, models.

The relationship between each pairing of tongue pressure and arch measurement variable appeared to be positive and reasonably linear (Fig 11). The data shows trends between discrete tongue pressure points, but data in between points is uncertain. Correlation estimates described the strength of these associations as follows: arch perimeter (r=0.99, CI: (0.50, 0.99)), anterior arch perimeter (r=0.99, CI: (0.75, 0.99)), arch length (r=0.98, CI: (0.25, 0.99)), and incisor spacing (r=0.99, CI: (0.75, 0.99)). However, it should be noted that although the interval for arch length does not include 0, it is still very wide, which raises questions about the magnitude of the true relationship. Intercanine width (r=0.95, CI: (-0.27, 99)) and intermolar width (r=0.96, CI: (-0.27, 0.99)) both have large magnitude correlations. However, due to the span of the intervals, confidence in both the true direction and true magnitude of the linear association between the independent and dependent variable is much lower. Despite this, the relationship between tongue pressure and both intercanine width and intermolar width appear visually to be positive and linear.

### Lip-Cheek Pressure Effects on Dental Displacement Patterns

A representative displacement vector plot of mandibular teeth resulting from lip-cheek pressure of 136.5 kg/m^2^ (mean literature value) is shown in Fig 12. As illustrated, lip-cheek pressure results in generalized inward, constrictive, crown movement. Surprisingly, the greatest displacements from lip-cheek pressure are found at the canines, premolars, and lateral incisors. Note that, for all teeth, a *distal* displacement component results for all crowns as the lip-cheeks apply pressure perpendicular to the arch. Qualitative examination of teeth displacements revealed that lip-cheek pressure results in minimal root movement compared to crown movement.

**Fig 12.**
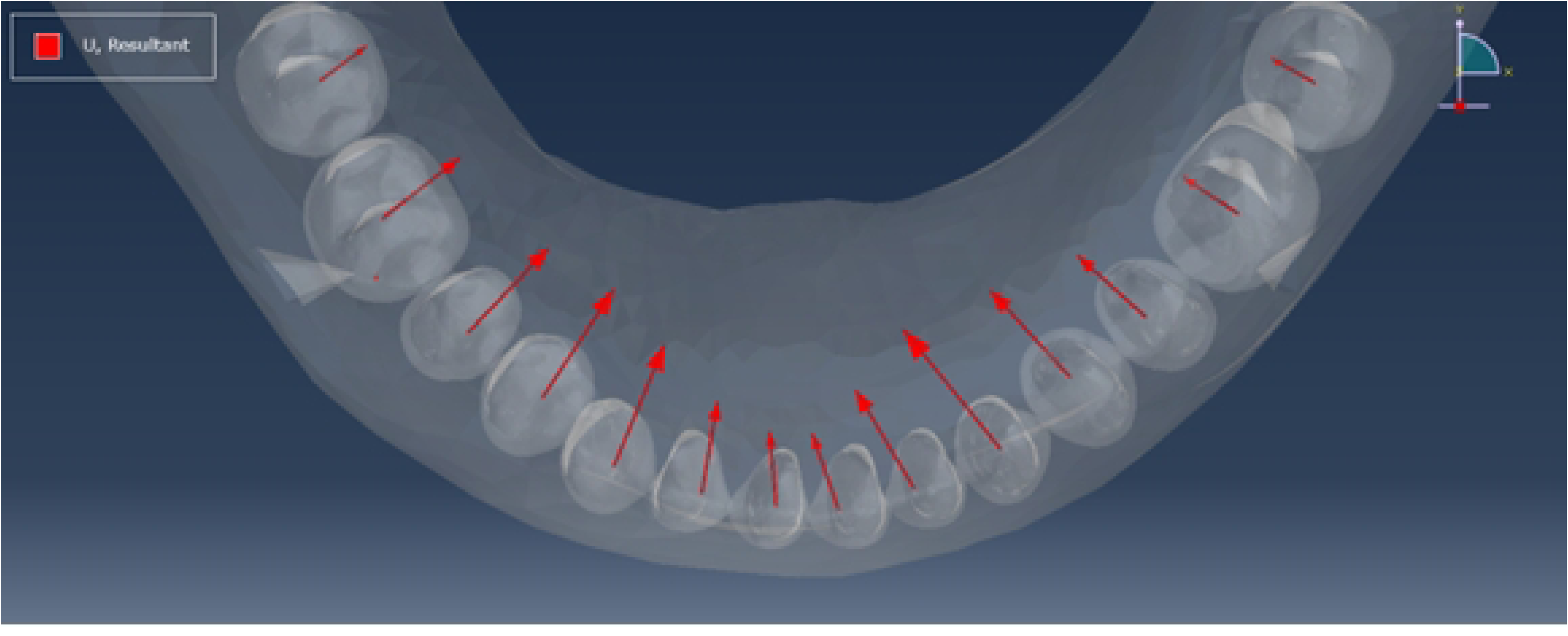
Scaled displacement vector plot of mandibular teeth crowns resulting from lip-cheek pressure of 136.5 kg/m2 (mean literature value).

An occlusal (X-Z plane) image of a lip-cheek pressure loaded mandibular arch model (328.3 kg/m^2^, high range value) is shown in Fig 13. Plots of corresponding arch measurements of unloaded and three loaded mandibular arches are provided in Fig 14. All results quantitatively corroborate the qualitative effects of lip-cheek pressure suggested in the vector plot above. That is, lip-cheek pressure results in significant anteroposterior and transverse arch constriction / collapse.

**Fig 13.**
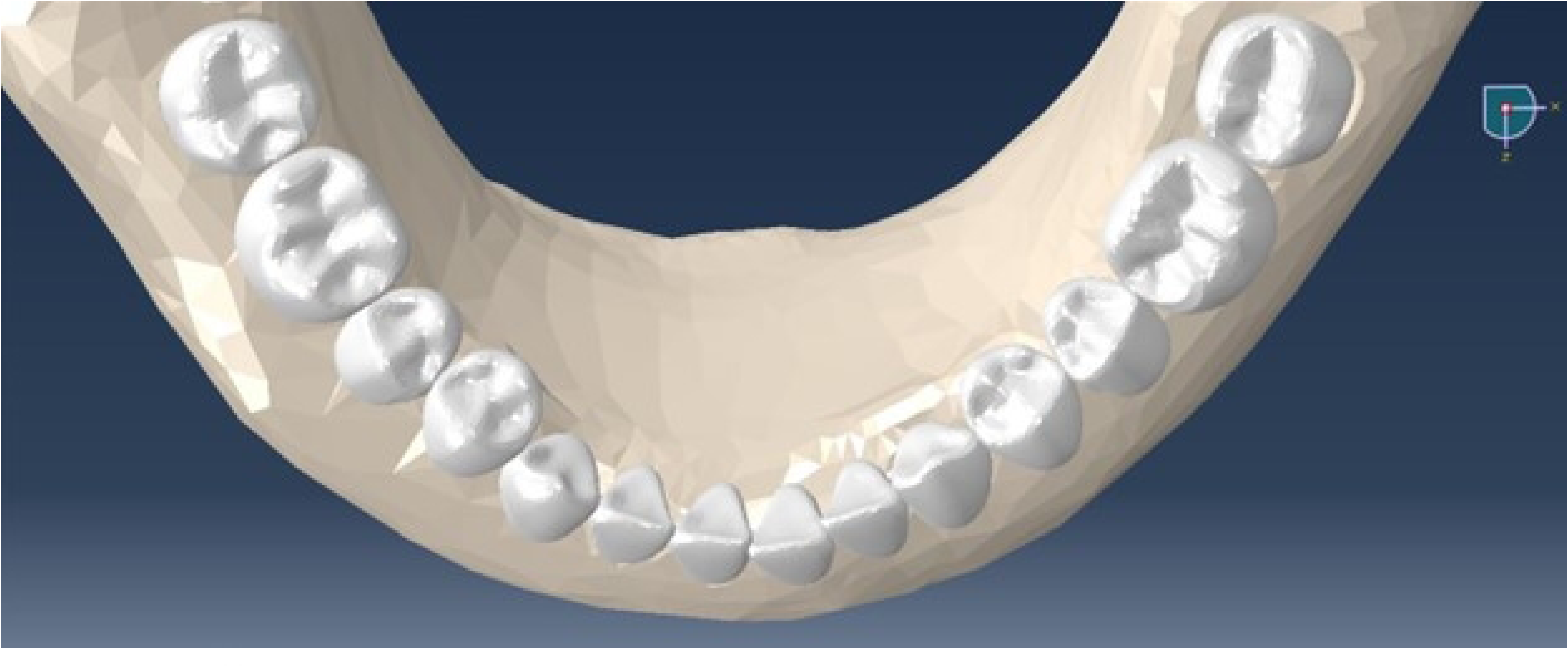
Mandibular arch loaded using lip-cheek pressure model (328.3 kg/m2, high range literature value).

**Fig 14.**
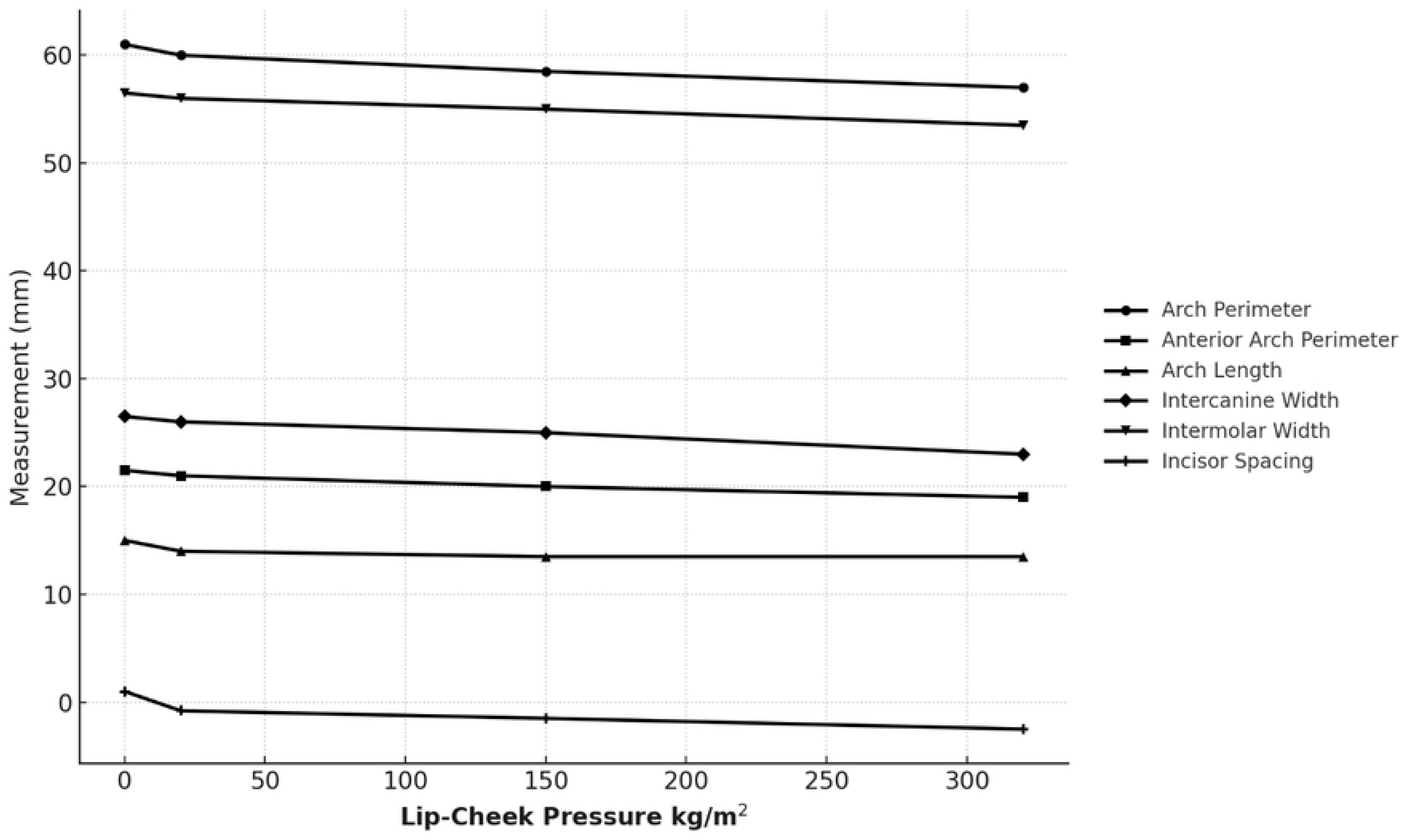
Effect of lip-cheek pressure (0-328.3 kg/m2) on dental arch parameters.

Under the influence of a mean (literature defined) lip-cheek pressure, mandibular arch perimeter decreased by 1.11 mm, or 1.8%, and the anterior arch perimeter decreased by 0.8 mm, or 3.7%. These changes are seen as incisor space closure / increased incisor crowding during loading.

Arch length reduction worsened with increasing lip-cheek pressure with the high value lip-cheek pressure model (328.3kg/m^2^) leading to a collapse in arch length of 1.37 mm, or 9%. Arch length reduction was due to greater distal displacement of anterior teeth and premolars compared to distal displacement of molars, a finding confirmed by evaluating reference node positions of teeth in unloaded and loaded models relative to mandibular encastre elements.

Intercanine and intermolar widths constricted under the influence of lip-cheek pressure. At mean lip-cheek pressure, the mandibular intercanine width decreased by 1.69 mm, or 6.3%, and the intermolar width decreased by 3.07mm, or 5.4%. An additional decrease in intercanine width by 1.63 mm, or 6.6%, and intermolar width by 1.51 mm, or 2.7%, occurred when the lip-cheek pressure increased from 136.5 kg/m^2^ to 328.3 kg/m^2^. Interestingly, slightly greater arch collapse occurred at the canines (3.32 mm, 13%) than at the first molars (3.07 mm, 5%) when lip-cheek pressures increased to the highest value. A marked increase in incisor crowding (2.55mm) resulted from mean lip-cheek pressure loading. Incisor crowding continued to increase under the highest value lip-cheek pressure.

The relationships between lip-cheek pressure and the dependent variables seem to be non-linear and logarithmically decreasing (Fig 14). The data shows trends between discrete lip-cheek pressure points, but data in between points is uncertain. Although for each relationship considered in isolation, the non-linear trend could be considered the result of random error in the observations, this is not the case when all relationships are considered overall. Because each pairing with lip-cheek pressure shows the same logarithmically decreasing trend, it is likely that this is the true shape of the relationships. Because of that, correlation estimates for arch perimeter (r=-0.97, CI: (-0.99, 0.14)), anterior arch perimeter (r=-0.94, CI: (-0.99, 0.38)), arch length (r=-0.81, CI: (-0.99, 0.78)), intercanine width (r=-0.99, CI: (-0.99, -0.40)), intermolar width (r=-0.99, CI: (-0.99, -0.43)), and incisor spacing (r=-0.84, CI: (-0.99, 0.74)) do not hold bearing even when the confidence intervals do not include 0. Regardless, visually, these relationships display a strong negative association, even if that association is non-linear.

### Interdental (Transseptal Fiber) Force Effects on Dental Displacement Patterns

A representative displacement vector plot of mandibular teeth resulting from an interdental (transseptal fiber) force of 0.0367 kg (mean literature literature) is shown in Fig 15. A generalized inward, constrictive crown movement is apparent, and the greatest inward displacement is at the canines. Note that, for anterior teeth and premolars, a *distal* component of displacement results from interdental tensional loading. However, both first and second molars are subjected to a *mesial* displacement. Qualitative evaluation of teeth displacements revealed that interdental force generated minimal root movement compared to crown movement.

**Fig 15.**
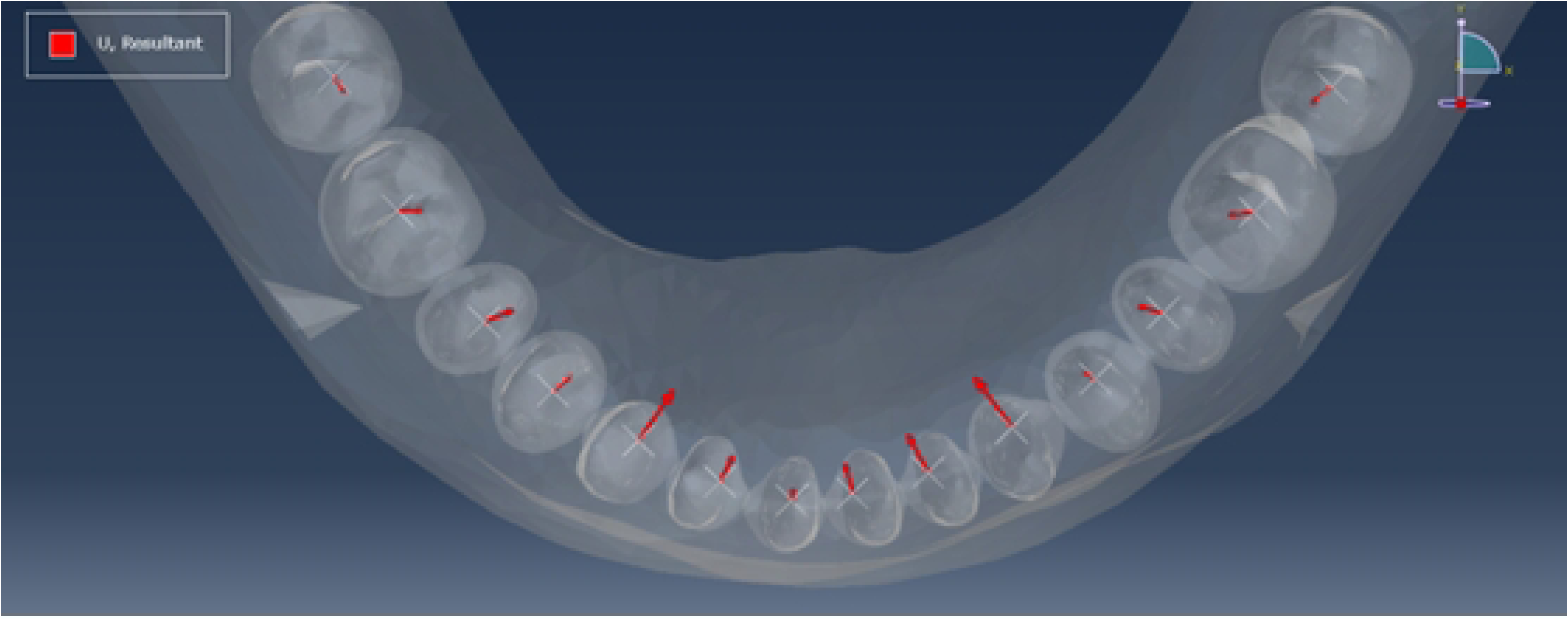
Scaled displacement vector plot of mandibular teeth crowns resulting from interdental (transseptal fiber) force of 0.0367 kg (mean literature value).

An occlusal (X-Z plane) image of an interdental (transseptal fiber force) loaded mandibular arch model (0.0869 kg, high range value) is shown in Fig 16. Plots of corresponding arch measurements are provided in Fig 17. All results quantitatively corroborate the effects of interdental force suggested in the vector plot above. That is, interdental force creates anteroposterior and transverse constriction and collapse.

**Fig 16.**
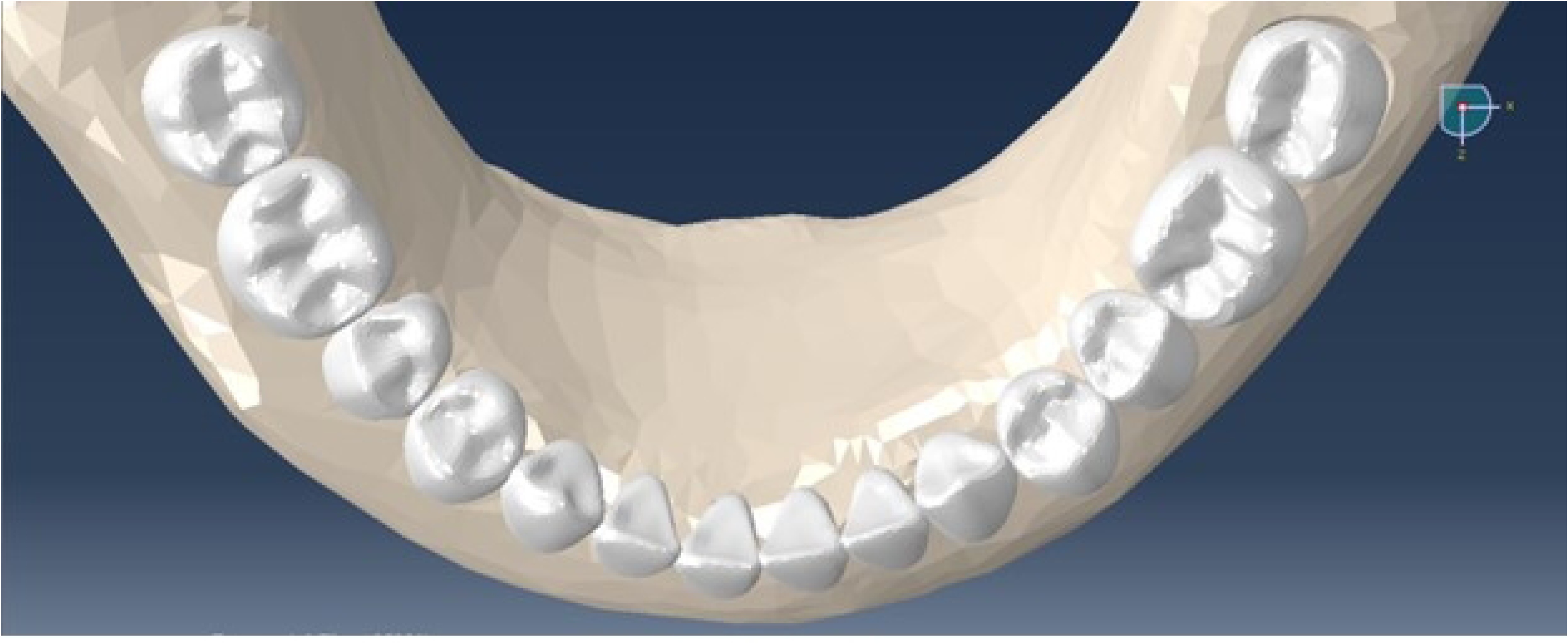
Mandibular arch loaded using interdental (transseptal fiber) force model (0.0869 kg, high range literature value).

**Fig 17.**
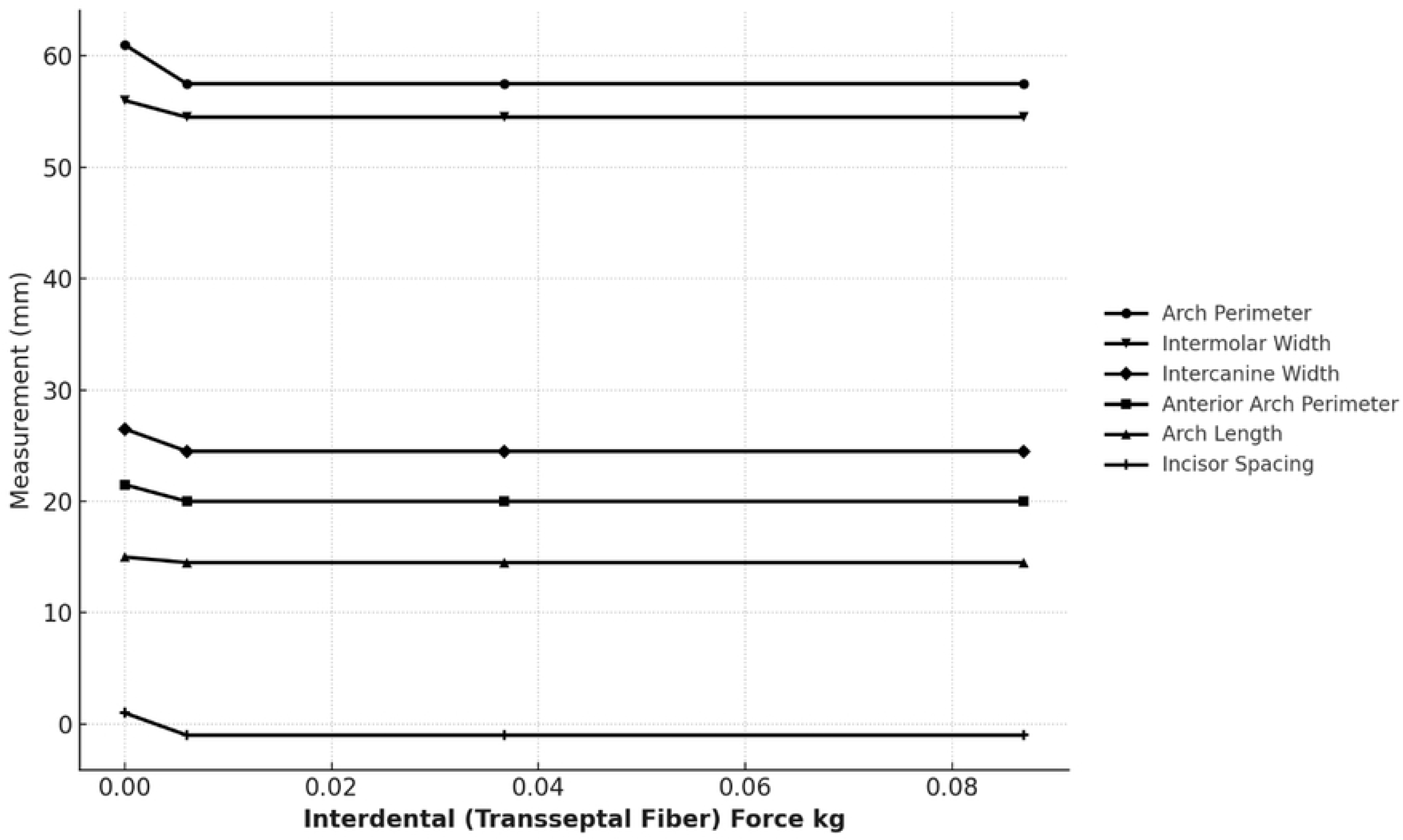
Effect of increasing interdental (transseptal fiber) force (0-0.0869 kg) on dental arch parameters.

Under the influence of a mean (literature defined) interdental force, mandibular arch perimeter decreased by 3.61mm, or 5.9%, and anterior arch perimeter decreased by 1.94 mm, or 9%. These changes are seen as incisor space closure / increased crowding during loading.

At the high range value of interdental force (0.0869 kg), the arch length decreased by 0.77 mm, or 5.1% due to mesial molar movement, space closure, and slight incisor lingual tipping movement (compare incisor uprightness in Fig 9 to Fig 16). Mesial molar movement was confirmed by determining molar reference node movement in unloaded and loaded models relative to mandibular encastre elements.

Intercanine and intermolar widths constricted. With mean interdental force loading, the mandibular intercanine width decreased by 2.63mm, or 9.9%, and the intermolar width decreased by 1.8mm, or 3.2%. Slightly greater constrictions were seen when the interdental loading increased to the high value.

Incisor crowding occurred with interdental loading. With a mean interdental load, incisor crowding increased from baseline (1.06 mm spacing) to 2.99 mm crowding. Incisor crowding decreased very slightly (by 0.015mm) when the interdental force was increased from the mean to high range value. Calculation of exact individual incisor node movements relative to encastre mandibular elements revealed that this very slight decrease in crowding was due to the method of spacing / crowding measurement - mesial slippage of the mandibular right central incisor mesial contact and consequent slight lengthening of the anterior perimeter measurement.

The relationships between transseptal fiber force and the dependent variables also appear to be logarithmically decreasing (Fig 17). The data shows trends between discrete interdental force points, but data in between points is uncertain. Compared to the cheek-lip pressure relationships, the transseptal fiber force relationships appear to have an even stronger non-linear association. Thus, correlation estimates between transseptal fiber force and arch perimeter (r=-0.51, CI: (-0.98, 0.87)), anterior arch perimeter (r=-0.53, CI: (-0.99, 0.86), arch length (r=-0.80, CI: (-0.99, 0.65)), intercanine width (r=-0.61, CI: (-0.99, 0.82)), intermolar width (r=-0.56, CI: (-0.99, 0.85), and incisor spacing (r=-0.48, CI: (-0.98, 0.88) are not an appropriate measure of the strength of these associations. However, visually the strength of these *negative* non-linear associations appears very strong.

### Dental Displacement Patterns Resulting from Simultaneous Tongue Pressure, Lip-Cheek Pressure, and Interdental Force Loading

A displacement vector plot of mandibular teeth resulting from simultaneous combined loading of 0.0366 kg (mean literature value) interdental force, 136.5 kg/m^2^ (mean literature value) lip/cheek pressure, and 100 kg/m^2^ (mean literature value) tongue pressure is shown in Fig 18. Generalized inward, constrictive, crown movement is apparent with the greatest inward displacement occurring at the canines. Note that, for anterior teeth and premolars, a *distal* component of displacement results from simultaneous application of these continuous, or quasi-continuous, loads. However, both first and second molars are subjected to a *mesial* displacement. Qualitative evaluation of teeth displacements revealed that simultaneous application of these three loads resulted in minimal root movement compared to crown movement.

**Fig 18.**
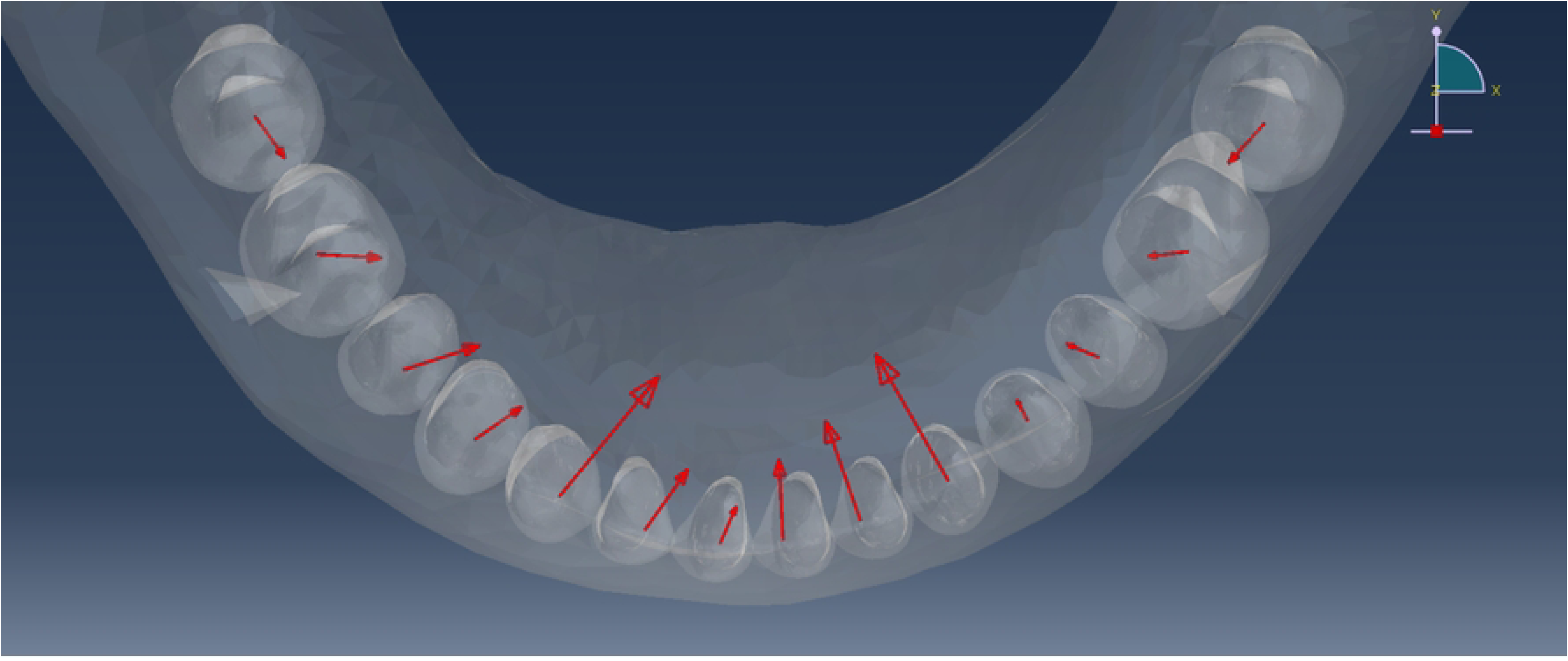
Displacement vector plot of mandibular teeth crowns resulting from simultaneous combined loading of inderdental force, lip-cheek pressure, and tongue pressure. Forces and pressures are literature mean values (0.0366 kg interdental force, 136.5 kg/m^2^ lip-cheek pressure, and 100 kg/m^2^ tongue pressure).

An occlusal (X-Z plane) image of a mandibular arch simultaneously loaded with 0.0366 kg (mean value) interdental force, 136.5 kg/m^2^ (mean value) lip-cheek pressure, and 100 kg/m^2^ (mean value) tongue pressure is shown in Figure 19. All results quantitatively corroborate the effects shown in the vector plot above, including anteroposterior and transverse arch constriction, arch collapse, and incisor space loss / increased crowding.

**Fig 19.**
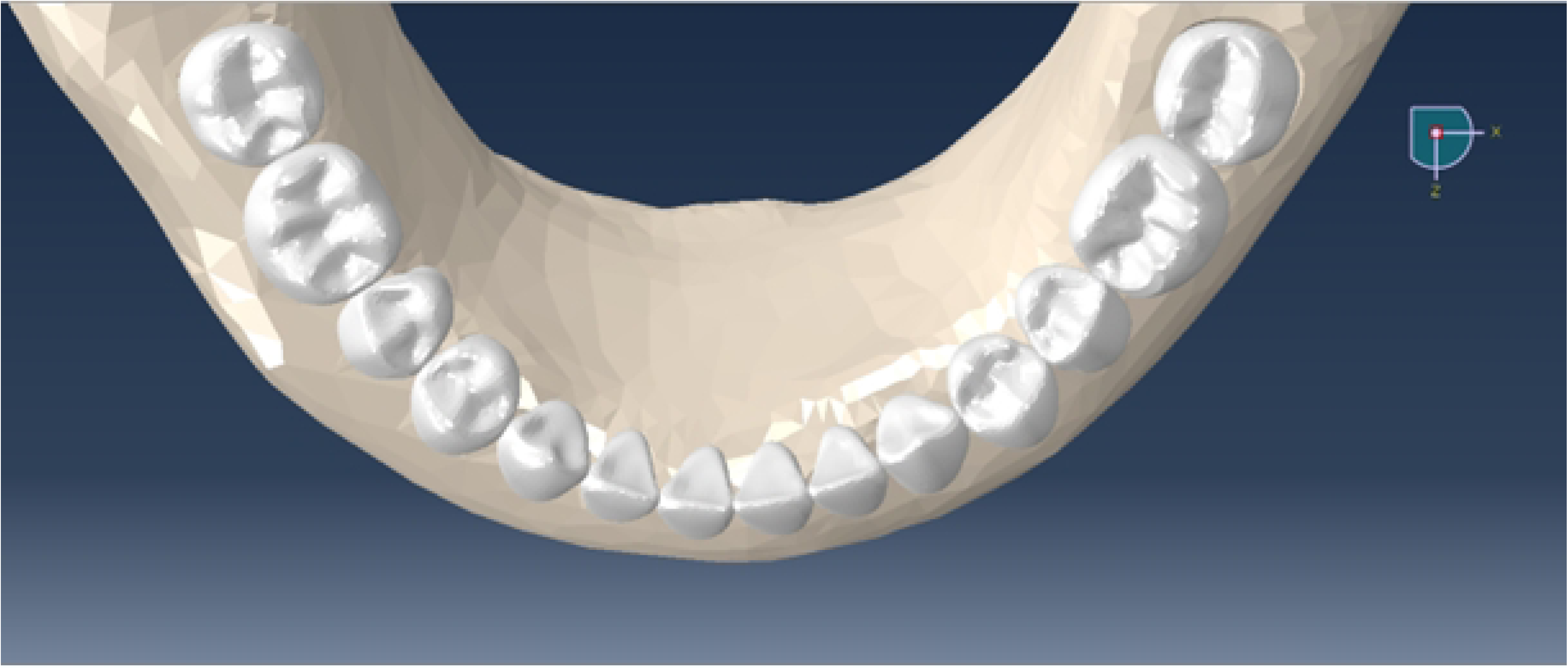
Mandibular arch simultaneously loaded with 0.0366 kg interdental force, 136.5 kg/m2 lip-cheek pressure, and 100 kg/m2 tongue pressure. All forces and pressures are mean literature values.

Under the influence of simultaneous mean tongue pressure, lip/cheek pressure, and interdental force, the mandibular arch perimeter decreased by 2.81mm, or 1.3%, and anterior arch perimeter decreased by 1.55mm, or 7.2%. These changes are seen as mild canine / incisor lingual collapse and crowding during loading (compare Fig 9 to Fig 19). Further, the arch length decreased by 0.88 mm, or 5.6% due to mesial molar movement, space closure, and incisor lingual tipping movement. Mesial molar movement was confirmed by determining molar reference node movement in unloaded and loaded models relative to mandibular encastre elements.

Intercanine and intermolar widths constricted with simultaneous loading. The mandibular intercanine width decreased by 1.85mm, or 6.9%, and the intermolar width decreased by 1.14mm, or 2.0%. When viewing the mandibular arch superimposition (Fig 20), lingual crown displacements of anterior teeth and premolars are readily seen – as are lingual and mesial molar crown movements. Finally, space loss occurred with simultaneous loading. Loss of space (increased crowding) worsened from baseline (1.06mm unloaded spacing reduced to 0.59mm spacing when loaded).

**Fig 20.**
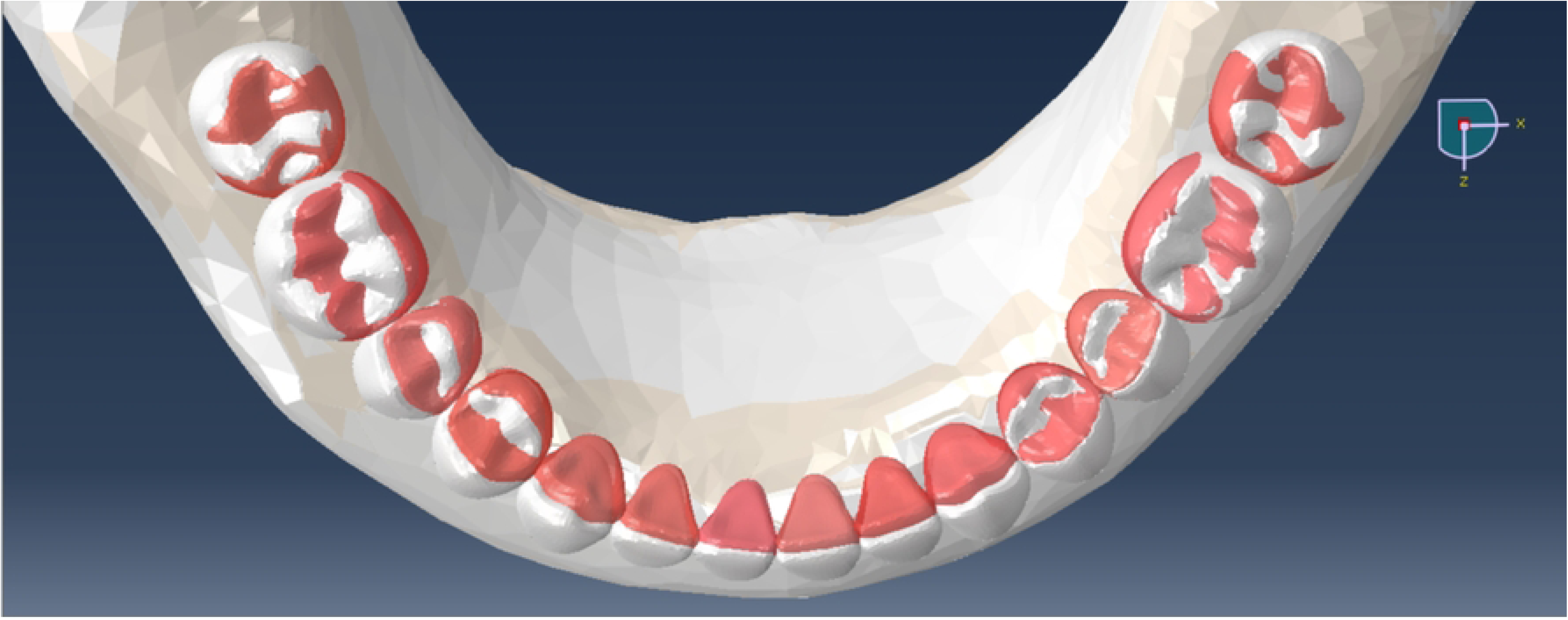
Superimposition of unloaded model (white teeth) and model simultaneously loaded (red teeth) with 0.0366 kg interdental force, 136.5 kg/m2 lip/cheek pressure, and 100 kg/m2 tongue pressure. All pressures and forces are mean literature values.

The principal findings of this preliminary study are that, hitherto undocumented, simultaneous application of (literature-defined average) lip-cheek pressure, tongue pressure, and interdental force to the mandibular dentition tends to collapse the mandibular anterior arch (intercanine width reduction / incisor crowding) and displace molars mesially (mesial molar drift). These novel findings mirror clinical reality in both treated and untreated patients and reflect this first attempt we are aware to explain the etiology of mandibular arch collapse through either modeling in vivo experimentation.

This study is important because a striking, unexplored knowledge gap exists in our specialty: an incomplete understanding of mandibular anterior arch collapse etiology. While significant advances have been made during the past century in orthodontic diagnosis and treatment, the equally important retention phase of care remains in a state of relative stasis, with lifelong mechanical retention being the only guarantee for post-treatment dental alignment. We hope that that this preliminary study establishes a foundation and serves as a vehicle encouraging other researchers to pursue this line of inquiry with the goal being shorter retention periods and stable dental alignment tailored to each patient’s individual needs.

Promising avenues for further study using this model as a starting point include the impact of overbite increase, mandibular growth, and the anterior component of occlusal force on mandibular anterior arch collapse and incisor crowding / alignment relapse. Additionally, the impact of variations in mandibular cortical and trabecular bone architecture, lip/cheek/tongue pressure variation with deflection, root anatomy variation, hyalinization and undermining resorption of varying thickness periodontal ligaments, ranges of bone elastic moduli, linear and non-linear bone response, and keratinized gingiva are all areas poised for exploration.

"If I have seen further, it is by standing on the shoulders of giants," is a phrase popularized by Issac Newton. We are fortunate today to have the technology to model and investigate the phenomena considered in the present study, but the foundation for this study was established decades ago by orthodontic giants including Dr. William Proffit who suggested that an equilibrium exists in the positioning of teeth in the arch and that the major primary factors in this dental position equilibrium may be resting tongue pressure, lip-cheek pressure, and forces created within the periodontal membrane.[5,6] The present study supports his notion by underscoring the importance of these three, relatively light but long-acting, intra-oral loads in dictating tooth position. Further, it demonstrates how the constrictive, collapsing effects of simulated lip-cheek pressure and interdental force is opposed by, and somewhat balanced by, the expansive effects of tongue pressure, with the former two dominating.

Other luminaries, including Drs. Robert Little [2,49], Peter Shapiro and Vincent Kokich [4] reported the phenomena of mandibular anterior arch collapse, mandibular intercanine width reduction, and increased mandibular anterior crowding over time, irrespective of orthodontic extraction treatment, serial extraction treatment, non-extraction treatment (generalized spacing), arch expansion, and even non-treatment. In untreated patients Little [49] reported that, over time, arch length and width decreased with mild to moderate crowding apparent into early adulthood, and untreated adults had a mean decrease of intercanine width over time. He noted that amongst all groups studied arch width and arch length constriction was a normal physiologic process occurring in both treated and untreated patients.[49] The present study supports his reported findings of increased incisor crowding and intercanine width reduction under simultaneous tongue pressure, lip-cheek pressure, and interdental force loading.

Further, the present study’s finding that simulated tongue pressure causes dental arch expansion supports the clinical observation reported when lip-cheek forces are removed during Frankel appliance wear. That is, by removing the effects of lip-cheek perioral musculature from the dentition, a shift in the equilibrium of intraoral forces occurs with tongue pressure producing measurable transverse expansion, particularly in the premolar and molar regions, and mandibular incisor advancement.[50,51]

In terms of validation, or how well our model captured the physical behavior of the real-world it simulates, the results qualitatively reveal that the model is valid. Simulated tongue pressure results in outward / expansive movement of the mandibular dental arch exactly as observed when cheek / lip pressures are removed by Frankel appliance buccal shields. [50,51] Likewise, simulated lip-cheek pressure results in inward tipping movement of mandibular anterior teeth precisely as observed when anterior tongue pressure is reduced / eliminated via treatment with bonded tongue spurs.[52] Finally, simulated interdental (transeptal fiber) force results in migration of mandibular teeth as demonstrated by Moss and Picton [21,27] plus mandibular anterior arch collapse, as suggested by Southard.[24] When combined, these three simulated loads create mandibular anterior arch collapse, inter-canine width collapse, and incisor crowding - phenomena reported extensively in the literature.[2–4]

With respect to our model’s representation of tooth movement, although significant strides have been made in our understanding of tooth movement biology in recent years, the detailed mechanisms of this mechanotransduction process, including mechanocoupling, biomechanical coupling, cell-to-cell signaling, and effector response remain to be elucidated. Areas requiring further exploration include periodontal ligament viscoelasticity, hyalinization formation and resorption, alveolar bone deformation behavior, and mechanical stress regulation of stem/progenitor cell differentiation into osteoblast and osteoclast lineages, among others.[36,41,53–57] Due to this incomplete understanding, all orthodontic tooth movement models are approximations, fall short of fully capturing the exceedingly complex and incompletely understood dynamic resorption and deposition processes that occur clinically, and must operate within the confines of model assumptions and model complexity limitations.

We considered various measures of mandibular incisor spacing / crowding prior to beginning this study. The method chosen was robust, permitting ready measurement of either increases in crowding or increases in spacing under load application. However, just as every cephalometric analysis suffers from one or more inherent weaknesses, the method we chose to measure crowding / spacing has a weakness. Specifically, we observed that under the greatest interdental force load, when increased incisor crowding occurred via central incisor midline contact *mesial* slippage, that this one specific movement increased the length of both left and right anterior arch perimeter arms, leading to the very slight decreased crowding measurement, despite the fact incisor crowding had increased.

Mesial migration of posterior teeth (mesial molar drift) is a well-recognized phenomenon in humans, although its etiology has never been fully understood.[25] One half century ago Moss and Picton [21] studied the possible cause of mesial migration by repeatedly removing posterior teeth contacts in *Macaca irus* monkeys which resulted in teeth being drawn toward the opened contact. Furthering their research, they isolated the transseptal fibers’ influence in *Macaca Irus* by removing cheek and tongue pressures with acrylic domes and surgically excising the transseptal fibers. They concluded that interdental fibers play a key role in mesial molar drift. [27,28] The results of the present study corroborate the phenomena of mesial molar displacement / molar drift under the influence of interdental force alone, but especially under the combined influence of simulated simultaneous interdental force, lip-cheek pressure, and tongue pressure loads where a clear mesial molar displacement / drift was evident.

Due to the shape of the mandibular arch, application of simulated tongue pressure in a direction perpendicular to the arch results in a mesial component of force applied to all teeth. Does the tongue play a role in mesial drift of mandibular molars? Or, as teeth wear interproximally with chewing, does the tongue hold anterior teeth forward while transeptal fibers (interdental forces) pull posterior teeth forward? These questions await further study.

In a similar fashion, simulated lip-cheek pressure applied perpendicular to the mandibular arch results in a distal displacement tendency for all teeth, even the molars, while simulated interdental force between teeth results in a distal component of displacement for anterior teeth and premolars, but a *mesial* displacement of first and second molars. For second molars the reason for mesial displacement was intuitively obvious since fiber pull was limited to distal pull from first molars alone. For first molars a mesial displacement component is still present but mitigated by distal fiber pull from second molars.

We were surprised to find simultaneous loading of tongue pressure, lip-cheek pressure, and interdental force resulted in the greatest dental displacement at the *canines* – where the arch turns the corner (Fig 18). This heightened canine displacement is reflected in worsening intercanine width reduction and lower incisor crowding. At the same time, incisors were displaced to the mesial and lingual, causing crowding independently. Molars and premolars collapsed towards the lingual and together.

When comparing upper range tongue pressure arch changes to upper range lip-cheek pressure arch changes, the amount of arch length *increase* from tongue pressure was outweighed by the arch length *decrease* from lip-cheek pressure. It was therefore not surprising to discover that the simultaneous application of mean tongue pressure, lip pressure, and interdental force resulted in a notable diminution in arch length. Such a phenomenon would yield a further tendency for mandibular arch collapse.

For FEA modeling to be helpful in understanding any biomechanical system, two major conditions should be met: 1) the objective of the simulation study must be to gain knowledge about the system that benefits clinical practice, and 2) the model itself must accurately represent the functioning of that system.[58] FEA modeling can help reveal how intraoral loads influence mandibular anterior arch collapse and direct us to further clinical study, perhaps leading our discipline to eventually modify retention protocol and improve long-term treatment stability.

With respect to the appropriateness of our model, the justification for study using FEA was found in two variables – load magnitude and load duration. In other words, lip-cheek pressure, tongue pressure, and interdental force are relatively light loads applied to the dentition over years, if not decades. Measuring in vivo effects of such intraoral light loads over extended periods of time is challenging, if not impossible, as evidenced by the lack of such clinical studies in the literature. FEA modeling, on the other hand, permitted ready study of light intraoral pressures and forces applied to the dentition irrespective of time. Further, FEA modeling permitted the effects of these loads to be studied individually or *in combination* – an endeavor many orthodontic researchers would consider clinically impossible.

It is important to realize that these findings of arch collapse, intercanine width reduction, and incisor crowding from simultaneous application of simulated tongue pressure, lip-cheek pressure, and interdental force is based upon the assumption of *continuous*, or quasi-continuous loading. Clinically, we would anticipate seeing patients who do not follow this pattern. Some patients may present with a lack of tongue posture (lack of tongue pressure) against the lingual of anterior teeth which would lead to greater arch collapse. Or some patients may exhibit tongue pressures much greater than the mean value tested herein which would overwhelm lip-cheek pressure and interdental force - leading to arch expansion / incisor spacing. Or some patients may present with taut lower lips and increased lip pressure against the anterior teeth which would lead to increased arch collapse. In other words, a wide range of individual variations in these loads and in their effects should be anticipated, in vivo.

## Conclusions

- The principal findings of this preliminary study are that, hitherto undocumented, simultaneous application of (literature-defined average) lip-cheek pressure, tongue pressure, and interdental force to the mandibular dentition tends to collapse the mandibular anterior arch (intercanine width reduction / incisor crowding) and displace molars mesially (mesial molar drift).
- Interdental (transeptal fiber) force and lip-cheek pressure collapse the mandibular anterior arch, decrease intercanine width, increase incisor crowding, and displace molars to the mesial.
- Tongue pressure expands the mandibular arch, increasing intercanine width, and generating inter-incisor spacing.
- These novel findings mirror clinical reality in treated and untreated patients, and this is the first attempt we are aware of explaining the etiology of mandibular arch collapse through modeling or clinical experimentation. We hope that that this study establishes a foundation and serves as a vehicle encouraging other researchers to pursue this line of inquiry with the goal being shorter retention periods and stable dental alignment tailored to each individual patient.

## Data Availability

N/A

## Acknowledgements

We thank Nicole Kallemeyn for her assistance with this project.

